# Dementia etiology classification using NULISA plasma biomarkers and machine learning

**DOI:** 10.64898/2025.12.02.25340706

**Authors:** Kelly N. DuBois, Subhamoy Pal, Amanda Cook Maher, Judith Heidebrink, Carol Persad, Bruno M. Giordani, Benjamin M. Hampstead, Kelly M. Bakulski, David G. Morgan, Nicholas M. Kanaan

## Abstract

**INTRODUCTION:** Accurate antemortem differentiation among dementia etiologies remains challenging, particularly for atypical or mixed clinical presentations. Multiplexed plasma proteomics paired with supervised machine-learning offers a minimally invasive and accessible approach for differential diagnosis.

**METHODS:** Plasma from 194 participants was analyzed using the NULISA CNS 120+ plasma biomarker panel. Differentially abundant protein patterns associated with AD, frontotemporal lobar degeneration, Lewy body disease, and vascular disease were identified. These features were used to train supervised XGBoost classifier models. Models were then applied to participants with mild cognitive impairment to generate data-driven predictions of etiology.

**RESULTS:** NULISA plasma biomarkers revealed disease-specific protein patterns. XGBoost classifiers differentiated disease etiologies with high specificity. Application of the models to participants with mild cognitive impairment yielded robust etiologic predictions.

**DISCUSSION:** These results support the feasibility of using multiplexed NULISA plasma proteomics, combined with machine learning, for differential diagnosis of complex neurodegenerative dementia etiologies.

**Highlights:**

- Multiplex plasma proteomics revealed distinct protein markers of dementia subtypes
- Supervised XGBoost classifiers accurately distinguished each dementia etiology
- Model application to unknown etiologies produced interpretable probability profiles
- The combined NULISA-machine learning framework demonstrates diagnostic feasibility

**Research in Context:**

1. Systematic review: The authors reviewed the literature using traditional sources (e.g. PubMed), meeting abstracts and presentations. NULISA technology has been utilized to analyze blood biomarkers in people with neurological diseases and differential protein expression based on clinical diagnosis and presumed etiologies has been observed. These citations are appropriately cited.
2. Interpretation: Our findings indicate that machine learning can be used in combination with NULISA technology to improve etiology prediction in people with dementia.
3. Future directions: Future studies using larger, more well-balanced participant cohorts will enable better understanding of the plasma biomarkers and demographic factors that best discriminate between dementia etiologies.

## 1. Background

Dementia encompasses many heterogeneous neurodegenerative and cerebrovascular syndromes with overlapping clinical features and underlying pathology [1,2]. Alzheimer’s disease (AD) remains the most common dementia worldwide, and it is expected to become more prevalent with aging populations [3]. Other etiologies such as frontotemporal lobar degeneration (FTLD), Lewy body disease (LBD) and vascular disease (VaD) together account for a substantial fraction of dementia cases and impose considerable disease burden on aging populations [4]. Early and accurate assessment of dementia etiology is essential for clinical trial enrollment and, with the advent of disease modifying therapies for AD, is increasingly crucial for clinical care decisions [5,6]. However, current clinical criteria have limited sensitivity and specificity to differentiate these disorders, particularly in atypical presentations or mixed pathologies [7].

Cerebrospinal fluid (CSF), Amyloid-β (Aβ) and tau positron emission tomography (PET) scans, and plasma biomarkers have significantly advanced AD diagnosis [8,9]. However, CSF collection is invasive, requiring lumbar puncture, which can be uncomfortable for participants and limits feasibility in large-scale or longitudinal studies. PET scans, while highly informative, are costly and require specialized equipment that is not widely available. Plasma-based biomarkers offer a more accessible, scalable and tolerable biofluid analysis. Plasma markers such as pTau-217, the Aβ42:40 ratio, and the pTau-217:Aβ42 ratio have aided discrimination of AD from other dementias [10–12]. This has led to improvements in understanding prognosis, clinical diagnosis, clinical trial participant stratification and identification of candidates for disease-modifying therapies [13].

Unbiased and multiplexed measurement of circulating plasma proteins offers a promising route to differential diagnosis of complex and overlapping dementia etiologies. Nucleic acid Linked Immuno-Sandwich Assay (NULISA) proteomics uses an antibody-based proximity ligation assay to offer ultrasensitive multiplexed measurements of plasma proteins, some of which reach sensitivity at attomolar concentrations [14]. The NULISA Central Nervous System (CNS) Disease Panel analyzes more than 120 proteins [15]. This panel accurately reflects known biomarkers of neurodegenerative disease and predicts Aβ PET status with performance consistent with other platforms [16–20].

Previously, it was demonstrated that this panel revealed condition-specific proteomic biomarkers across neurodegenerative disorders [20–22].

We leveraged NULISA proteomic data to validate the identification of etiology-specific proteins in participants enrolled in the Michigan Alzheimer’s Disease Research Center (MADRC) University of Michigan Memory and Aging Project (UM-MAP) cohort [23]. We selected 194 participants based primarily on presumed etiology. We first compared cognitively unimpaired (CU), AD, FTLD, LBD, and VaD participants to identify differentially abundant proteins. We then used these proteins to train supervised machine learning classifier models (XGBoost) to distinguish each etiology from the remainder of the cohort. XGBoost, a tree-boosting system, was chosen for its speed and scalability, regularization to prevent overfitting, and its ability to maintain performance with data missingness and multicollinearity [24]. Finally, we applied these derived models to participants with mild cognitive impairment (MCI), including those with variable clinical symptom history and fluctuating diagnosed etiology history, to generate data-driven predictions of etiology. Our results demonstrate that NULISA plasma biomarkers represent a scalable, minimally invasive framework to augment clinical assessment and improve antemortem diagnostic confidence.

## 2. Methods

### 2.1 Study Participants

A total of 194 UM-MAP (MADRC) participants were included (Table 1). MADRC recruitment sources comprised University of Michigan Health System clinics, the Wayne State University Healthier Black Elders Center, and community outreach. Written informed consent (with assent as appropriate) was obtained from participants or legally authorized representatives. Clinical procedures consisted of a clinician-performed neurological examination, standardized neuropsychological testing by trained staff, venous blood sampling for biomarker analyses, and neuroimaging (for a subset of participants). The protocol complied with federal and state regulations and received approval from the University of Michigan IRB (HUM00000382); compensation was provided per IRB approval.

**Table 1:** Study participant characteristics by clinical phenotype. Numeric variables are summarized using median (25th percentile, 75th percentile). Categorical variables are summarized using number (percent). Note: *p-*values for continuous variables are from Kruskal-Wallis rank sum tests p-value and for categorical variables from Pearson’s Chi-squared test and adjusted *p-*value with Bonferroni corrections for multiple comparisons. Abbreviations: CDR-Sum, Clinical Dementia Rating-Sum of Boxes; MoCA, Montreal Cognitive Assessment; CU, cognitively unimpaired; MCI, mild cognitive impairment; AD, Alzheimer’s disease; FTLD, frontotemporal lobar degeneration; LBD, Lewy body disease; VaD, vascular disease.

| Characteristics | Overall<br>(N=194) | CU<br>(N=35) | MCI<br>(N=53) | AD<br>(N=33) | FTLD<br>(N=30) | LBD<br>(N=26) | VaD<br>(N=17) | *p*-value | Adjusted<br>*p*-value |
| --- | --- | --- | --- | --- | --- | --- | --- | --- | --- |
| Age (years) | 72.0<br>(67.3,<br>77.0) | 73.0<br>(70.0,<br>76.0) | 72.0<br>(69.0,<br>77.0) | 70.0<br>(66.0,<br>76.0) | 64.0<br>(61.3,<br>68.8) | 77.0<br>(71.5,<br>80.0) | 76.0<br>(71.0,<br>82.0) | <0.001*** | <0.001*** |
| Sex (male) | 100<br>(51.5%) | 15<br>(42.9%) | 29<br>(54.7%) | 16<br>(48.5%) | 16<br>(53.3%) | 19<br>(73.1%) | 5<br>(29.4%) | 0.088 | 0.876 |
| Education (years) | 16<br>(14, 18) | 18<br>(15, 18) | 16<br>(14, 18) | 16<br>(23, 18) | 16<br>(14, 17.3) | 16<br>(14, 18) | 14<br>(14, 16) | 0.090 | 0.904 |
| Missing | 6 | 0 | 4 | 0 | 2 | 0 | 0 |  |  |
| APOE-ε4 allele<br>(any) | 88<br>(45.4%) | 8<br>(22.9%) | 23<br>(43.4%) | 22<br>(66.7%) | 10<br>(33.3%) | 7<br>(26.9%) | 8<br>(47.1%) | 0.052 | 0.524 |
| Missing | 13 | 0 | 7 | 1 | 3 | 1 | 1 |  |  |
| Race |  |  |  |  |  |  |  |  |  |
| White | 138<br>(71.1%) | 23<br>(65.7%) | 34<br>(64.2%) | 23<br>(69.7%) | 30<br>(100%) | 22<br>(84.6%) | 6<br>(35.3%) | <0.001*** | 0.004** |
| Black/African<br>American | 55<br>(28.4%) | 12<br>(34.3%) | 19<br>(35.8%) | 9<br>(28.1%) | 0<br>(0%) | 4<br>(15.4%) | 11<br>(64.7%) |  |  |
| CDR-Sum | 1.0<br>(0.6, 3.9) | 0<br>(0.0, 0.0) | 0.5<br>(0.5, 1.0) | 4.5<br>(2.5, 5.0) | 4<br>(1.6, 6.0) | 5.5<br>(2.0-8.9) | 0.5<br>(0.5, 1.0) | <0.001*** | <0.001*** |
| MoCA Total | 23<br>(18, 26) | 28<br>(26, 29) | 24<br>(22, 25) | 17<br>(12, 19) | 19<br>(11, 22) | 21<br>(15, 25) | 24<br>(21, 26) | <0.001*** | <0.001*** |
| Missing | 7 | 0 | 4 | 1 | 1 | 1 | 0 |  |  |
| Aβ PET |  |  |  |  |  |  |  |  |  |
| Positive | 39 | 0 | 18 | 20 | 0 | 1 | 0 | <0.001*** | <0.001*** |
| Negative | 28 | 15 | 11 | 0 | 1 | 0 | 1 |  |  |
| Missing | 127 | 20 | 24 | 13 | 29 | 25 | 16 |  |  |

### 2.2 Participant Clinical Characterization

Measurement of cognitive function was obtained using the Uniform Data Set (UDS) version 3 from the National Alzheimer’s Coordinating Center (NACC) [25,26]. This included the Clinical Dementia Rating scale (CDR) [27] and the Montreal Cognitive Assessment (MoCA) [28] used in this study.

Diagnosed etiologies were established for each participant through a consensus conference that included at least three MADRC clinicians (neurologists/neuropsychologists) after review of all visit materials. Eligible participants for this study were those clinically categorized at or near the date of blood collection as one of the following: 1) cognitively unimpaired (CU); 2) non-amnestic or amnestic, single-domain or multi-domain MCI of unknown or presumed AD etiology, which is referred to collectively as MCI; 3) amnestic multi-domain dementia syndrome consistent with Alzheimer’s disease etiology (AD); 4) cognitive impairment with a presumed etiology of frontotemporal lobar degeneration (FTLD); 5) cognitive impairment with a presumed etiology of Lewy Body disease (LBD); or 6) cognitive impairment with a presumed etiology of vascular brain injury (vascular disease, VaD). Also of interest were participants with variable clinical symptom history and fluctuating diagnosed etiology history, defined as 3 or more different diagnoses in their longitudinal clinical history that did not follow the progression of CU to MCI to a dementia.

Global cognition was characterized using the total MoCA score (range: 0-30) since it is a screening tool for cognitive impairment that assesses various aspects of cognitive functioning including visuospatial abilities, executive function, short-term memory, language, and orientation to time and place [29,30]. Higher MoCA scores indicate better cognitive functioning. The CDR is a clinical rating scale that gathers information from the participant and study partner to evaluate six aspects of cognitive and behavioral functioning (memory, orientation, judgment and problem-solving, community affairs, home and hobbies, and personal care). Two summary scores are calculated from the CDR [31,32]. We used the CDR Sum of Boxes (CDR-SB; range 0-18) score, since it provides more information and is better able to assess dementia gradations than the Global Score [33]. Lower CDR-SB scores indicate better cognitive functioning. All cognitive measures were performed by trained and certified Michigan ADRC staff.

Participant demographics [age in years, sex (male/female), race and ethnicity (NHW or non-Hispanic Black/African American, B/AA), educational attainment (years of education), and medical history] were ascertained at the clinical examination by the individual and/or a care partner-report. Aβ PET positivity was determined using PET Pittsburgh Compound B (PiB) amyloid imaging scan results [34]. Classification followed Centiloid criteria [35]: participants with Centiloid values of < 20 were considered Aβ–, while those with Centiloid values of ≥ 20 were classified as Aβ+. *APOE* genotyping was performed at the National Centralized Repository for Alzheimer’s Disease and Related Dementias by DNA isolation and analysis of two *APOE* single nucleotide polymorphisms (rs429358 and rs7412) using the buffy coat from blood samples. Either one or two copies of the *APOE*-ε4 allele was operationalized as the presence of *APOE*-ε4.

### 2.3 Plasma Biomarker Measurements

Participants’ blood was collected into 10 mL K2-ethylenediaminetetraacetic acid (EDTA) tubes. Plasma, red blood cells, and buffy coat were then processed and stored at −80°C until use. Samples underwent no more than 2 freeze/thaw cycles prior to use in biomarker assays.

#### 2.3.1 Simoa biomarker measurements

Plasma Aβ40, Aβ42, pTau-217, pTau-231, GFAP, NfL, TDP-43, sTREM2, and PlGF were measured on the Quanterix HD-X analyzer using the Simoa® Neurology 4-Plex E Advantage Plus (#104543 GFAP, NfL, Aβ42, Aβ40), Simoa® pTau-217 Advantage Plus (#104570), Simoa® pTau-231 Advantage Plus (#104512), Simoa® TDP43 (#103293), Simoa® sTREM2 Advantage Plus (#104543), and Simoa® PlGF Discovery (#102318) kits. After thawing and mixing, plasma samples were centrifuged at 4°C for 5 minutes at 10,000 x g. Supernatants were diluted according to manufacturer’s instructions using the instrument’s onboard dilution protocol and duplicates were run from a single well each on a 96-well plate. Eight-point calibration curves and sample measurements were determined on Simoa Analyzer software using a weighting factor 1/Y2 and a four-parameter logistic curve fitting algorithm. Two levels of quality control material were included in each batch. Two bridge samples were included with each batch to monitor inter-run assay reproducibility.

#### 2.3.2 NULISA biomarker measurements

Plasma samples from all participants were analyzed with the NULISAseq™ (Nucleic acid Linked Immuno-Sandwich Assay) platform CNS Disease panel 120 [15], including Aβ peptides, p-tau forms, NfL (or “NEFL” as from the panel nomenclature), and other markers of neurodegeneration, inflammation, and vascular health (**Table S1**). Sample and data analysis was performed according to kit manufacturer protocols, including log2 transformation of the data and NULISA Protein Quantification (NPQ) on the logarithmic scale (Alamar Biosciences). Specifically, data are first normalized using internal and inter-plate controls. The data are then rescaled by multiplication with a factor of 10^4^. After this, +1 is added to all values. The data are then log2 transformed to make the data more symmetrical, forming a more normal distribution and accounting for the effects of outliers in subsequent statistical analyses. These values are defined as NPQ units, which are on a logarithmic scale. Differences in NPQ can be interpreted as log2 (fold change). NULISA markers were detected with high mean sensitivity across the three plates containing samples (95.4%, 91.6%, and 93.1%, respectively, range 85%-100%) and with a low mean intraplate coefficient of variance (4.4%, 5.6%, and 6.9%, respectively). Only one sample (LBD group) had a high internal control coefficient of variance (109.4%), but upon further inspection had NPQ values that were similar in magnitude to other samples in the same clinical phenotype group, so the sample was retained for analysis.

### 2.4 Descriptive and Statistical Analyses

GraphPad Prism for macOS (version 10.5.0), R Statistical Software (version 4.5.0), and Python (version 3.12.11) were used for generating graphs and statistical comparisons. Q-Q plots and the Shapiro-Wilk tests were used to determine the normality of data. Due to a lack of normal distributions within the data sets, we described the distributions of continuous variables using median and quartiles. We described distributions of categorical covariates using count and frequency. Potential differences in demographic and clinical data by cognitive status, sex, or race and ethnicity were assessed with Chi-square and Mann-Whitney tests. We used scatter plots with median and interquartile range whiskers to visualize differences in biomarker levels between groups. Kruskal-Wallis tests followed by Dunn’s test for multiple comparisons were conducted to examine differences in biomarker levels across groups stratified by clinical phenotype. Correlations between continuous variables were calculated using Spearman correlation. For these descriptive tests, we reported *p*-values and Bonferroni multiple comparison adjusted *p*-values.

To test for differential protein expression between clinical phenotype group categories, we used linear regression models in the limma package [36] with NPQ units (unadjusted for covariates). Results were displayed with volcano plots using color schemes to highlight the markers that were significantly different between groups, considering uncorrected *p*-values or after false discovery rate (FDR) correction for multiple comparisons.

### 2.5 Supervised Machine Learning multivariate modeling

#### 2.5.1 Model training

We used five-fold cross-validation with training and testing sets to develop predictive models for participant groups. For each target diagnosed etiology (AD, FTLD, LBD, VaD), we constructed a one-versus-rest binary classification model using an imbalanced-learn pipeline [37]. We included only those participants in the CU group who were Aβ PET negative and participants in the AD group who were Aβ PET positive to increase reliability of these reference groups in the training cohort.

Participants with a clinical diagnosis of MCI, including those with a fluctuating longitudinal consensus diagnosis, were excluded from the training dataset and instead comprised the prediction dataset.

Candidate model features were identified from the differential-expression (limma) analyses; each participant group was compared to each of the others, and the features with the smallest *p*-values across these contrasts were the candidate biomarkers selected for each model. A tunable p-value cutoff (0.001-0.2) was optimally tuned for each one-versus-rest model to optimize the number of biomarkers used for prediction. To address class imbalance (underrepresentation of the target etiology (minority class) vs. the rest of the cohort) within each cross-validation fold, the minority class was up-sampled using synthetic minority oversampling technique (SMOTE) [38] with the number of neighbors set to 3. This setting ensured that each synthetic sample was created by interpolating between a real minority-class case and one of its three nearest minority neighbors, while keeping the number of generated samples sufficient to balance the class distributions.

The core classifier for each target etiology was an eXtreme gradient-boosted (XGBoost) decision tree model (XGBoost gradient-boosted decision tree (objective = binary:logistic, evaluation metric = log-loss) with tree_method=“hist” and random_state=42 to ensure reproducibility [24]. A stratified grid search within five-fold cross-validation tuned the number of trees (n_estimators), learning rate, tree depth, subsampling fraction, column-sampling fraction, and the limma p-value threshold.

#### 2.5.2 Model calibration and evaluation

For each diagnosis, the best cross-validated pipeline was further calibrated by isotonic regression [39,40] applied through the *CalibratedClassifierCV* from the Scikit-learn package [41,42]. Model performance was evaluated using out-of-fold predictions and summarized by balanced accuracy, F1 score, sensitivity, specificity, and area under the receiver operating characteristic curve (ROC AUC). A non-parametric bootstrap procedure was applied to the out-of-fold predicted probabilities to calculate the 95% confidence interval for each AUC.

2.5.3 Feature importance

To assess the relative contribution of individual proteins and clinical covariates, we extracted mean absolute SHapley Additive exPlanation (SHAP) values from the final fitted models [43]. Feature importance tables were exported for each diagnostic model to support downstream biological interpretation.

## 3. Results

### 3.1 Full Cohort Characteristics

Participants and associated demographic and clinical characteristics are described in **Table 1**. Significant differences were seen in the age of participants and the racial composition across clinical phenotype categories. In our non-AD cognitively impaired groups (FTLD, LBD, and VaD), participants in FTLD or LBD groups scored more similarly to those in the AD group on cognitive scores (CDR-Sum and MoCA Total) than those in the VaD group, who scored more similarly to those with MCI.

### 3.2 NULISA measurements compared to Simoa measurements

For many participants, plasma biomarkers (i.e. pTau-217, pTau-231, GFAP, NfL, TREM2, TDP43, PlGF, Aβ40, and Aβ42) were measured on the same blood draw using both Simoa and NULISA assays. We compared these measurements and found that all measurements of the same analyte on the two platforms correlated significantly, except for Aβ40 (**Figure S1**). Kruskal-Wallis tests demonstrated differences between participant groups for established biomarkers of neurodegeneration measured by NULISA, further underscoring the accuracy and clinical relevance of these measures (**Figure S2**).

### 3.3 Group comparisons of NULISA biomarker protein differential abundance

We set out to use the panel of NULISA plasma biomarker measurements to train predictive models that could predict the best matched etiology for individual MCI participants. Our model training cohort excluded individuals with a clinical diagnosis of MCI and those with a variable longitudinal clinical diagnostic and etiological histories. We also included only those participants in the CU group who were Aβ PET negative and participants in the AD group who were Aβ PET positive to increase reliability of these reference groups in the training cohort. Clinical and demographic characteristics of the 109 participants that comprised the training cohort are described in **Table 2**.

**Table 2:**
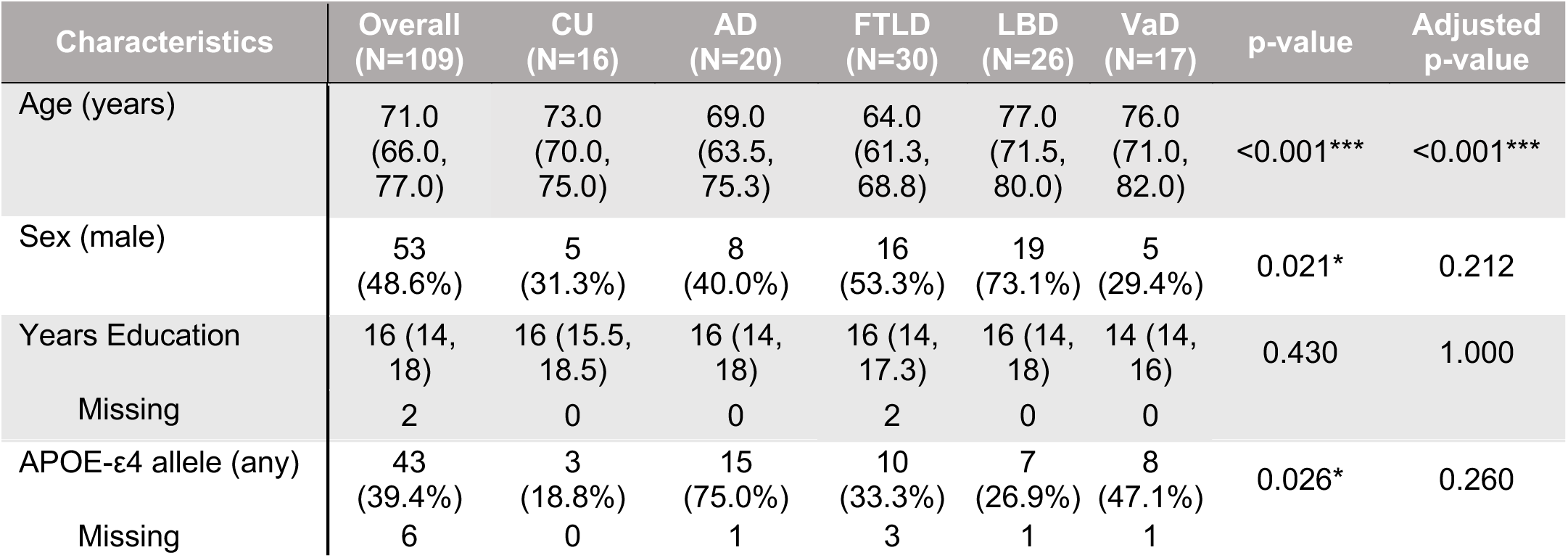
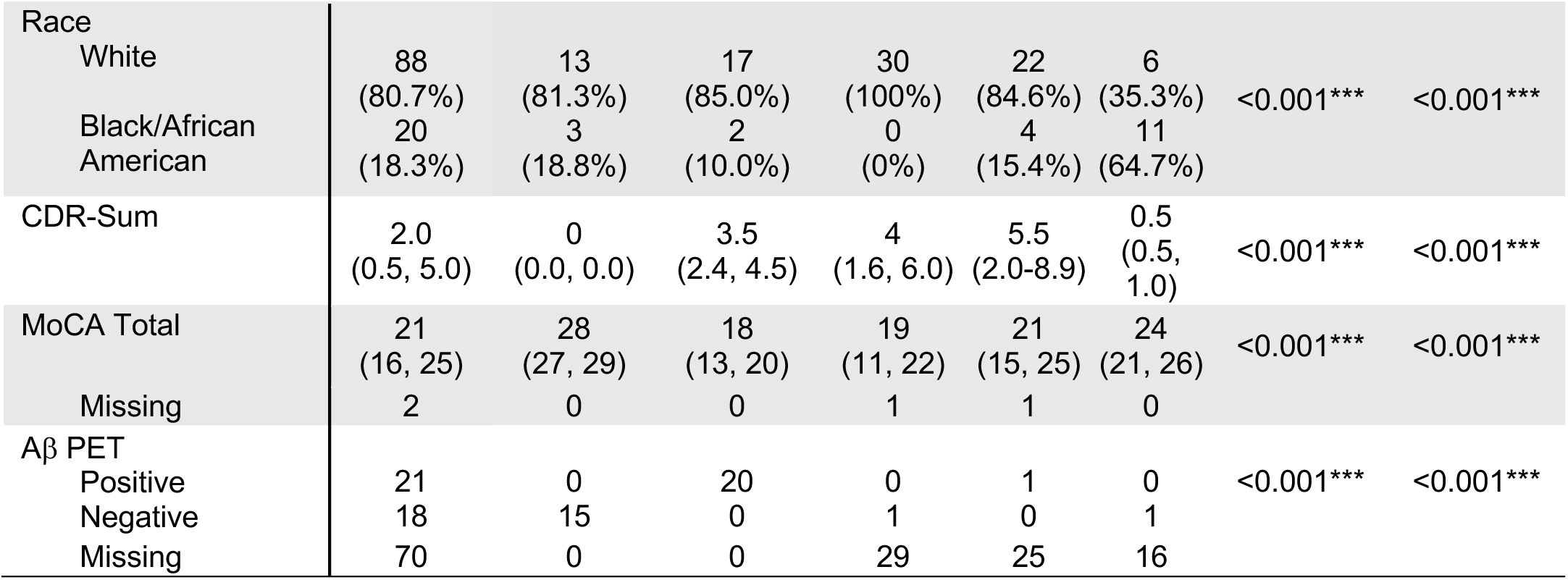
Training cohort characteristics by etiology group. Numeric variables are summarized using median (25th percentile, 75th percentile). Categorical variables are summarized using number (percent). Note: *p-*values for continuous variables are from Kruskal-Wallis rank sum tests p-value and for categorical variables from Pearson’s Chi-squared test and adjusted *P* value with Bonferroni corrections for multiple comparisons. Abbreviations: CDR-Sum, Clinical Dementia Rating-Sum of Boxes; MoCA, Montreal Cognitive Assessment; CU, cognitively unimpaired; MCI, mild cognitive impairment; AD, Alzheimer’s disease; FTLD, frontotemporal lobar degeneration; LBD, Lewy body disease; VaD, vascular disease.

Prior to prediction modeling, we assessed differential abundance of NULISA plamsa proteins using limma analysis. Using this cohort, we first compared Aβ PET positive AD participants with Aβ PET negative CU participants. Proteins associated with AD and neurodegeneration were significantly increased in Aβ PET positive AD participants, such as phosphorylated Tau species and GFAP (**Figure 1A**). In addition, BACE1, NPTX1, and VGF were significantly increased, while CRP, PDGFRB, and CXCL8 were significantly decreased in Aβ PET positive AD participants compared to Aβ PET negative CU participants (**Figure 1A**). Next, we compared CU, MCI, and AD participants.

**Figure 1:**
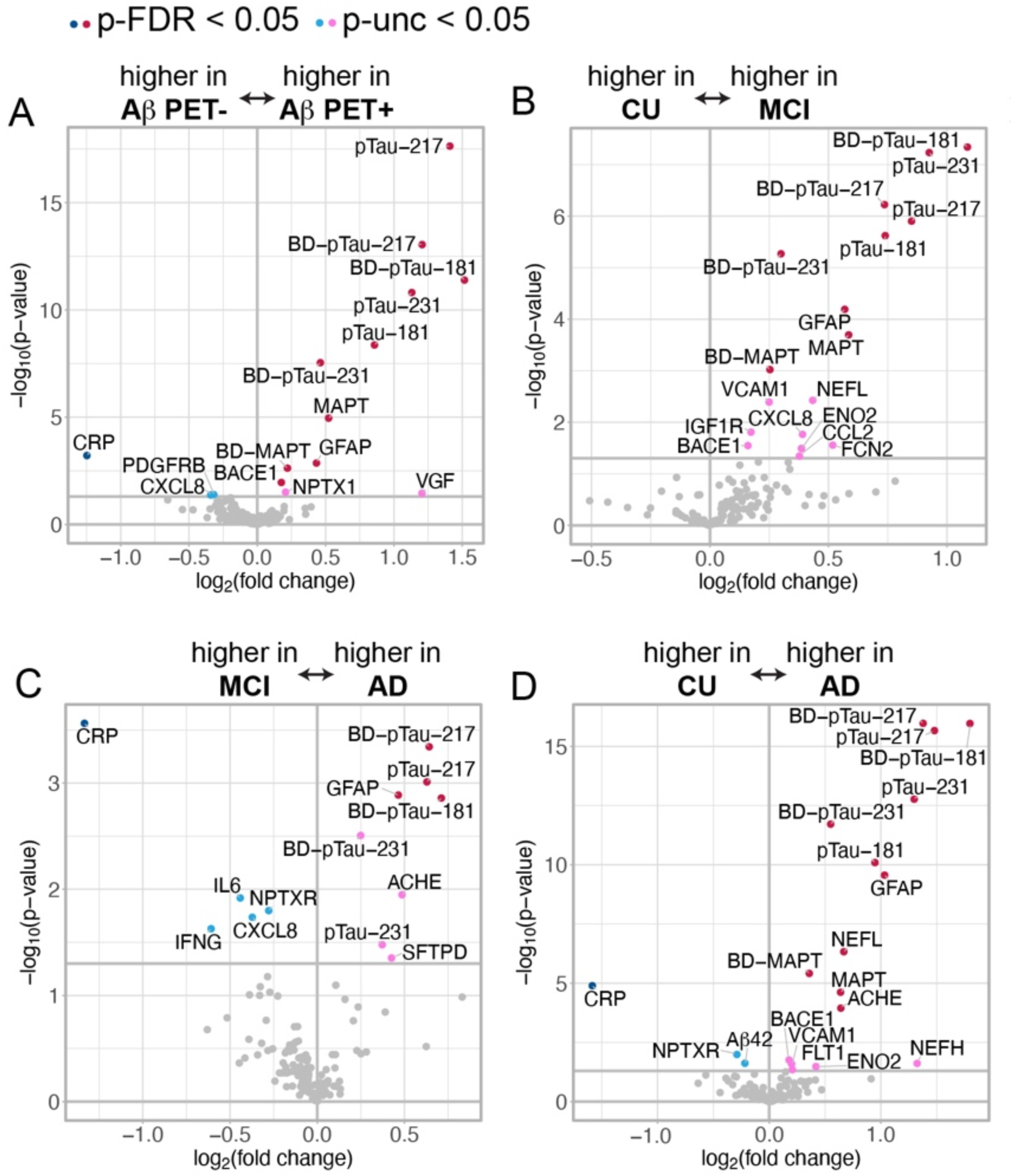
Comparisons between groups on NULISA CNS panel biomarkers. (A) Aβ PET positive compared to Aβ PET negative participants. (B) Participants with a clinical diagnosis of mild cognitive impairment (MCI) compared to cognitively unimpaired (CU) participants. (C) AD participants compared to MCI participants. (D) AD participants compared to CU participants. For each comparison, the panel on the right represents proteins that are increased compared to the reference group and the panel on the left represents proteins that are decreased compared to the reference group. Darker dots (red and blue) represent comparisons with p<0.05 corrected for multiple comparisons (false discovery rate correction, FDR) while lighter dots (pink and light blue) represent comparisons with uncorrected (unc) p<0.05. The gray vertical line signifies a fold change of 0, the gray horizontal line signifies a -log10 *p*-value of 0.05 (uncorrected).

Phosphorylated and total tau species and GFAP were increased in MCI compared to CU and further escalated in AD compared to MCI (**Figures 1B, 1C**). Interestingly, CXCL8 was increased in MCI compared to CU but decreased in AD compared to MCI, suggesting that it may be associated with MCI of a non-AD etiology (**Figures 1B, 1C**). IL6, IFNG, SFTPD, FCN2, CCL2, and IGF1R were also differentially regulated in MCI compared to CU or AD (**Figures 1B, 1C**). VCAM1, NfL (NEFL), ENO2, and BACE1 were higher in both MCI and AD participants compared to CU (**Figures 1B, 1D**). When comparing AD to CU, the additional elevated proteins were NEFH, FLT1 and ACHE, while CRP, NPTXR, and Aβ42 were lower (**Figure 1D**).

To identify protein profiles unique to non-AD cognitively impaired groups (FTLD, LBD, and VaD), we compared each of these clinical phenotype groups to Aβ PET negative CU participants (**Figure 2**) or to Aβ PET positive AD participants (**Figure 3**). While many of the same protein biomarkers that were increased in AD were increased in non-AD groups compared to CU, such as phosphorylated tau and GFAP, several unique proteins were significantly increased. For example, plasma NfL/NEFL was the most elevated protein when comparing FTLD, LBD, and VAD to CU participants (**Figure 2A-C**). Aβ species were differentially abundant in FTLD and VaD compared to CU participants. Aβ42 was decreased in FTLD and Aβ38 and Aβ40 were increased in VaD (**Figure 2A,C**). ICAM1 and CXCL1 were increased in VaD compared to both CU (**Figure 2C**) and AD (**Figure 3C**). VCAM1 was increased in VaD compared to CU participants (**Figure 2C**). In FTLD, CHIT1 and CCL11 were decreased compared to CU (**Figure 2A**) and AD participants (**Figure 3A**). Generally, non-AD groups had decreased levels of phosphorylated tau and GFAP compared to AD (**Figure 3**). Aβ species were more prevalent in LBD and VaD compared to AD (**Figure 3B,C**).

**Figure 2:**
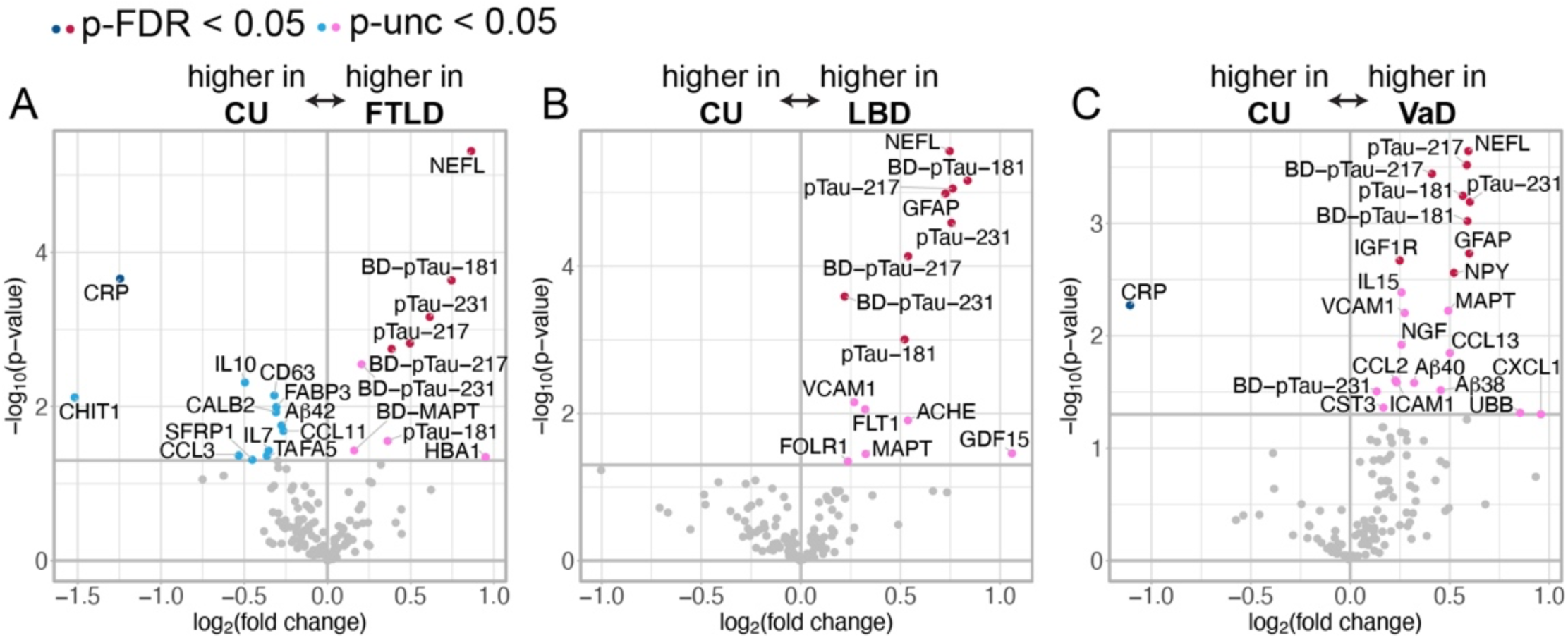
Comparisons between frontotemporal lobar degeneration (FTLD), Lewy body disease (LBD) or vascular disease (VaD) and cognitively unimpaired (CU) participants on NULISA CNS panel biomarkers. (A) FTLD participants compared to CU participants. (B) LBD participants compared to CU participants. (C) VaD participants compared to CU participants. For each comparison, the panel on the right represents proteins that are increased compared to the reference group and the panel on the left represents proteins that are decreased compared to the reference group. Darker dots (red and blue) represent comparisons with p<0.05 corrected for multiple comparisons (false discovery rate correction, FDR) while lighter dots (pink and light blue) represent comparisons with uncorrected (unc) p<0.05. The gray vertical line signifies a fold change of 0, the gray horizontal line signifies a -log10 *p*-value of 0.05 (uncorrected).

**Figure 3:**
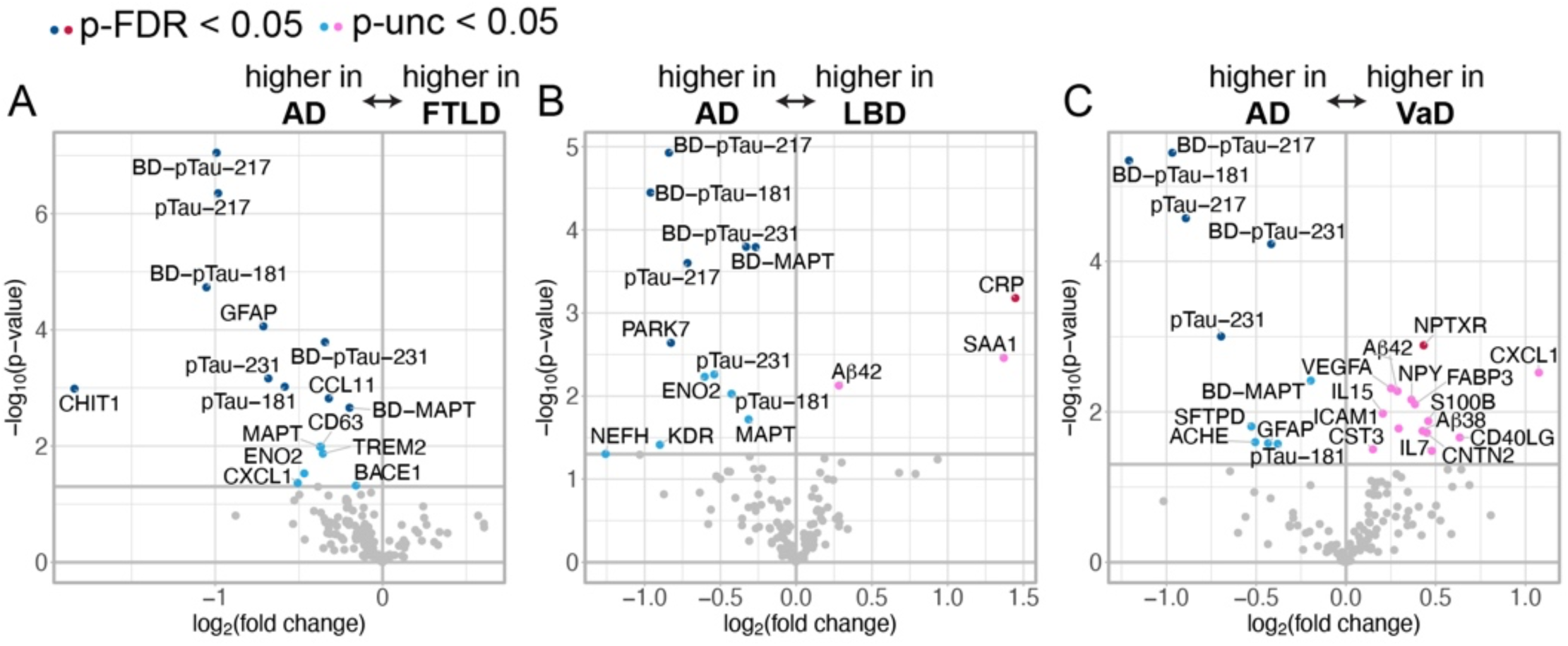
Comparisons between frontotemporal lobar degeneration (FTLD), Lewy body disease (LBD) or vascular disease (VaD) and AD participants on NULISA CNS panel biomarkers. (A) FTLD participants compared to AD participants. (B) LBD participants compared to AD participants. (C) VaD participants compared to AD participants. For each comparison, the panel on the right represents proteins that are increased compared to the reference group and the panel on the left represents proteins that are decreased compared to the reference group. Darker dots (red and blue) represent comparisons with p<0.05 corrected for multiple comparisons (false discovery rate correction, FDR) while lighter dots (pink and light blue) represent comparisons with uncorrected (unc) p<0.05. The gray vertical line signifies a fold change of 0, the gray horizontal line signifies a -log10 *p*-value of 0.05 (uncorrected).

Non-AD groups were also compared with each other. CXCL1 was lower in FTLD compared to LBD and VaD and lower in LBD compared to VaD (**Figure 4**). Aβ species were differentially abundant, with levels of Aβ40, Aβ42, and Aβ38 all being lower in FTLD than in VaD, levels of Aβ40 and Aβ42 lower in FTLD than LBD, and levels of Aβ38 lower in LBD than in VaD (**Figure 4**). CD63, CCL17, ICAM1, FABP3, NPY, CCL3, IGF1R, and IL15 were all decreased in FTLD and LBD compared to VaD (**Figure 4**). HBA1 was increased in FTLD compared to LBD and VaD (**Figure 4**).

**Figure 4:**
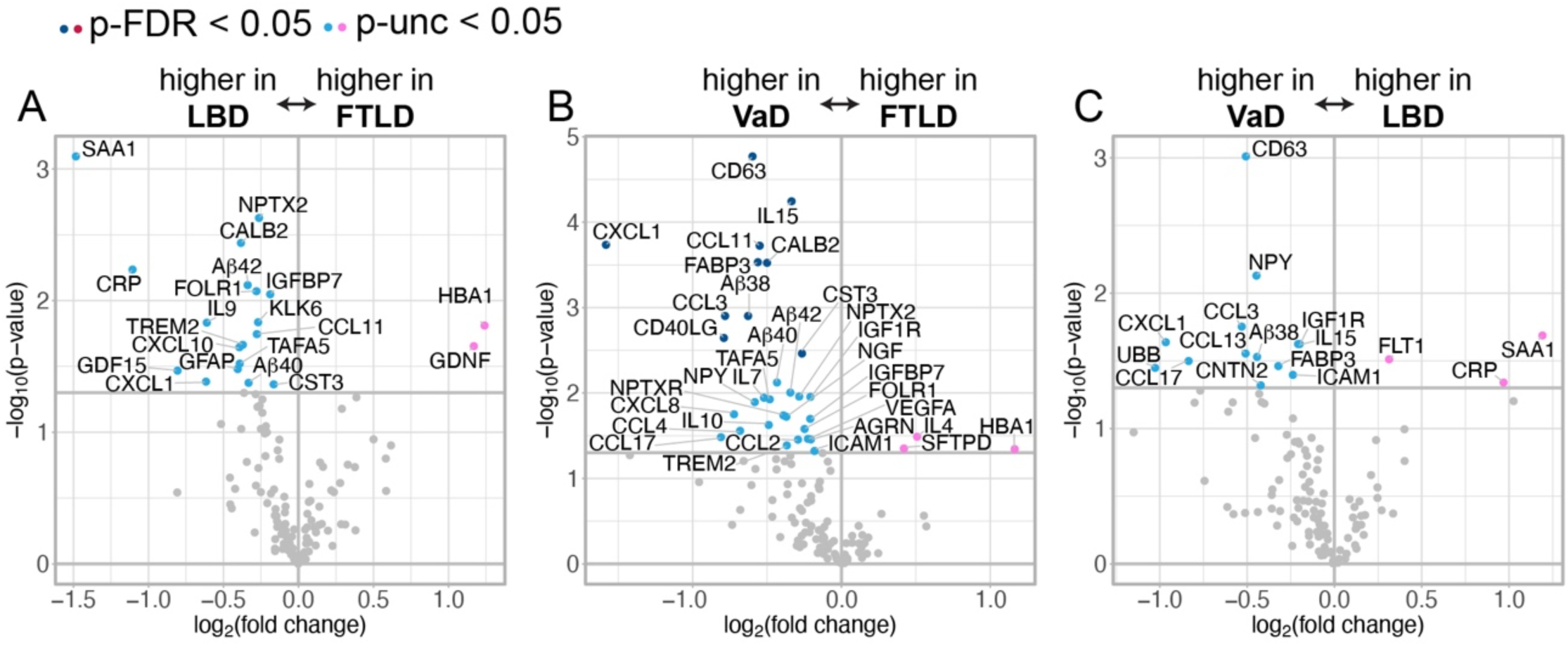
Comparisons between frontotemporal lobar degeneration (FTLD), Lewy body disease (LBD) and vascular disease (VaD) on NULISA CNS panel biomarkers. A, FTLD participants with a compared to LBD participants. B, FTLD participants compared to VaD participants. C, LBD participants compared to VaD participants. For each comparison, the proteins in the upper right quadrant represents those that are significantly increased compared to the reference group and the proteins in the upper left quadrant represents those that are significantly decreased compared to the reference group. Darker dots (blue) represent comparisons with p<0.05 corrected for multiple comparisons (false discovery rate correction, FDR), while lighter dots (pink and light blue) represent comparisons with uncorrected (unc) p<0.05. The gray vertical line signifies a fold change of 0, the gray horizontal line signifies a -log10 *p*-value of 0.05 (uncorrected).

### 3.4 Machine learning multivariable modeling of the association between biomarkers and participant group

Having established etiology-related differential protein abundance, we next determined whether the NULISA biomarker protein data could generate effective models for etiological predictions. Using our model training cohort (**Table 2**), we used five-fold cross validation to train one-versus-rest predictive models for AD, FTLD, LBD, and VaD, using NULISA plasma biomarker measurements, unadjusted for covariates. Out-of-fold predictions were used to calculate balanced accuracy, area under the receiver operating characteristic curve (ROC AUC, **Figure 5A**), F1 score, sensitivity, and specificity (**Table 3**).

**Figure 5:**
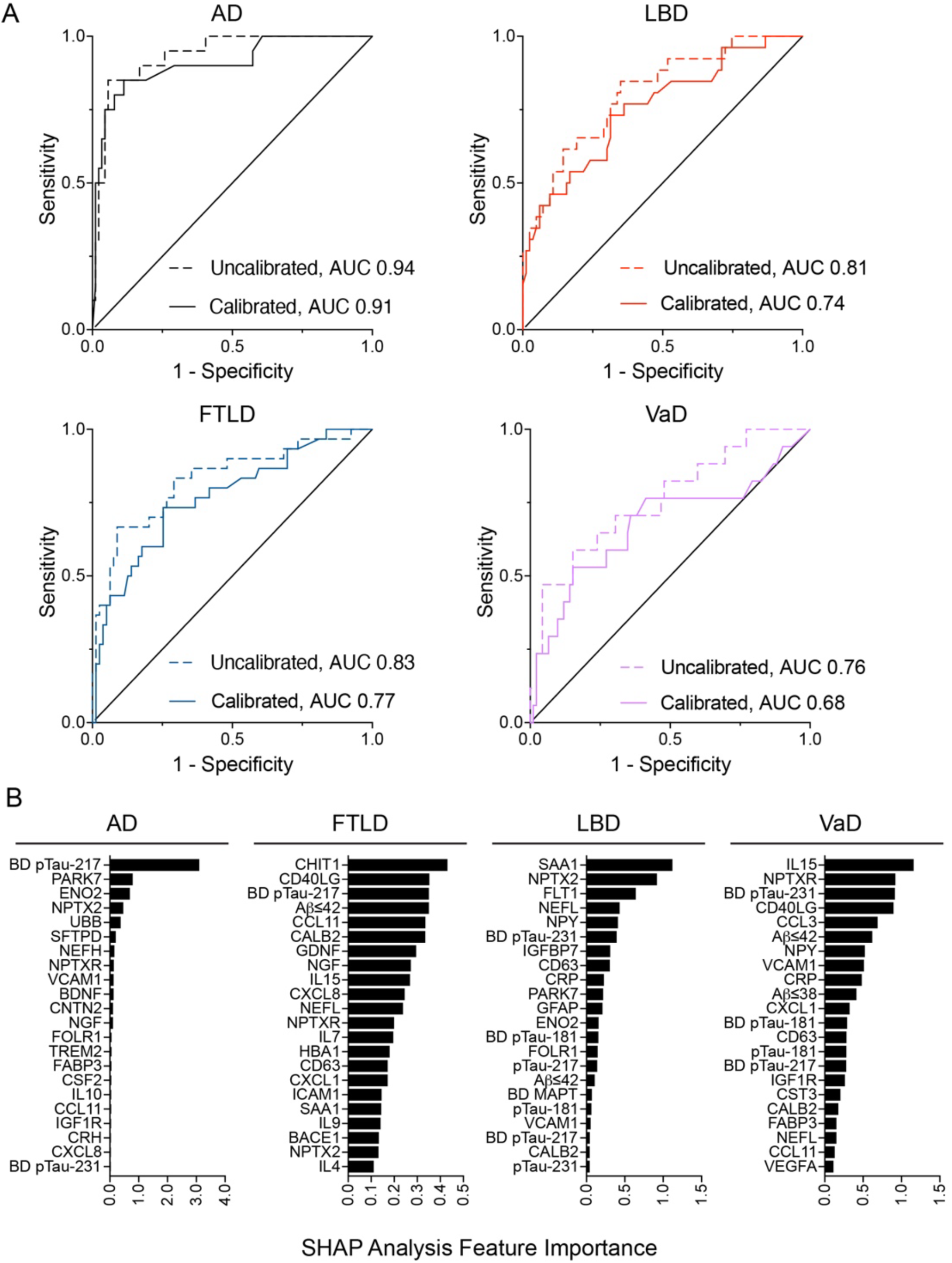
Trained XGBoost model classification discrimination and important model features. (A) Receiver operator characteristic curve area under the curve (AUC) for each model before (Uncalibrated) and after (Calibrated) calibration by isotonic regression comparing each participant group with the rest of the training cohort. (B) SHAP analysis of the top 22 contributing features to each one-versus-rest model. Abbreviations: AD, Alzheimer’s disease; FTLD, frontotemporal lobar degeneration; LBD, Lewy body disease; VaD, vascular disease. Full protein names can be found in **Table S1**.

**Table 3:**
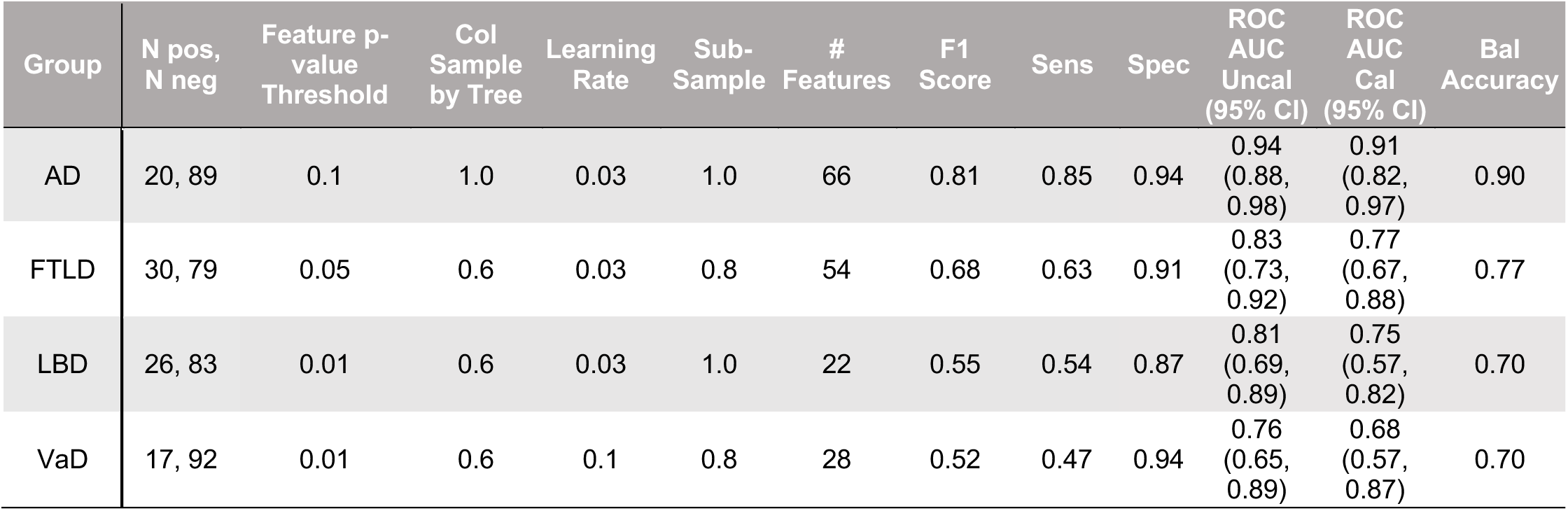
Model parameters and metrics for one-versus-rest model out of fold predictions shown for each final model by participant group. XGBoost max depth of 3 and XGBoost max number of estimators of 300 were chosen for every model. Abbreviations: AD, Alzheimer’s disease; FTLD, frontotemporal lobar degeneration; LBD, Lewy body disease; VaD, vascular disease; pos, positive; neg, negative; Col sample by tree, fraction of features used for building each tree; Sens, sensitivity; Spec, specificity; ROC AUC, receiver operator characteristic curve area under the curve; Uncal, uncalibrated; Cal, calibrated; Bal, balanced; 95% CI, 95% Confidence Interval.

Overall, this approach achieved robust discrimination of each dementia subtype, with AUCs ranging from 0.68 for VaD to 0.91 for AD in the calibrated models (**Figure 5A**). All models demonstrated higher sensitivity than specificity, indicating that they are more effective at identifying true positives than true negatives (**Table 3**).

SHapley Additive exPlanations (SHAP) analysis was used to quantify magnitude of the contribution of each plasma biomarker to each model (**Figure 5B**). Brain-derived pTau-217 (BD pTau-217) was the top feature for the AD model (**Figure 5B**). Chitinase 1 (CHIT1) was the largest contributor to the FTLD model, followed closely by CD40 ligand (CD40LG), BD pTau-217, and Aβ42 (**Figure 5B**). Serum amyloid A1 (SAA1), Neuronal pentraxin 2 (NPTX2), fms related receptor tyrosine kinase 1 (FLT1), and NfL/NEFL were important features for the LBD model (**Figure 5B**). The VaD model relied most heavily on interleukin 15 (IL15), neuronal pentraxin receptor (NPTXR), Brain-derived pTau-231 (BD pTau-231) and CD40LG (**Figure 5B**). BD pTau-217 was used as a feature in every model, underscoring its ability to discriminate among all dementia subtypes under investigation (**Figure 5B**). Some NULISA biomarkers, such as NfL/NEFL, Aβ42, calbindin 2 (CALB2), CD63 were in the top 22 features of all non-AD dementia models, demonstrating an ability to differentiate AD from other etiologies (**Figure 5B**). Abundance differences between participant groups for the top eight contributing NULISA biomarkers for each model and each participant group were plotted for the entire cohort (**Figure S3**).

### 3.5 Application: Etiology prediction for participants with MCI using NULISA biomarker measurements

We applied the final models to a set of 58 cognitively impaired participants from the UM-MAP cohort as our prediction cohort. All had a clinical diagnosis of MCI at least once in their MADRC visit history. For 27 participants, multiple longitudinal blood samples were included in the NULISA analysis, allowing us to examine etiology predictions over time. Plasma pTau-217 measurements (using Simoa) were available for 37 of the 58 MCI participants in this cohort. pTau-217 had a median value of 0.38 with an interquartile range of 0.26, 0.85. The relatively low pTau-217 plasma concentration in these cognitively impaired participants was expected to increase the likelihood of non-AD etiologies. Clinical and demographic characteristics of this prediction cohort are described in **Table 4**.

**Table 4:**
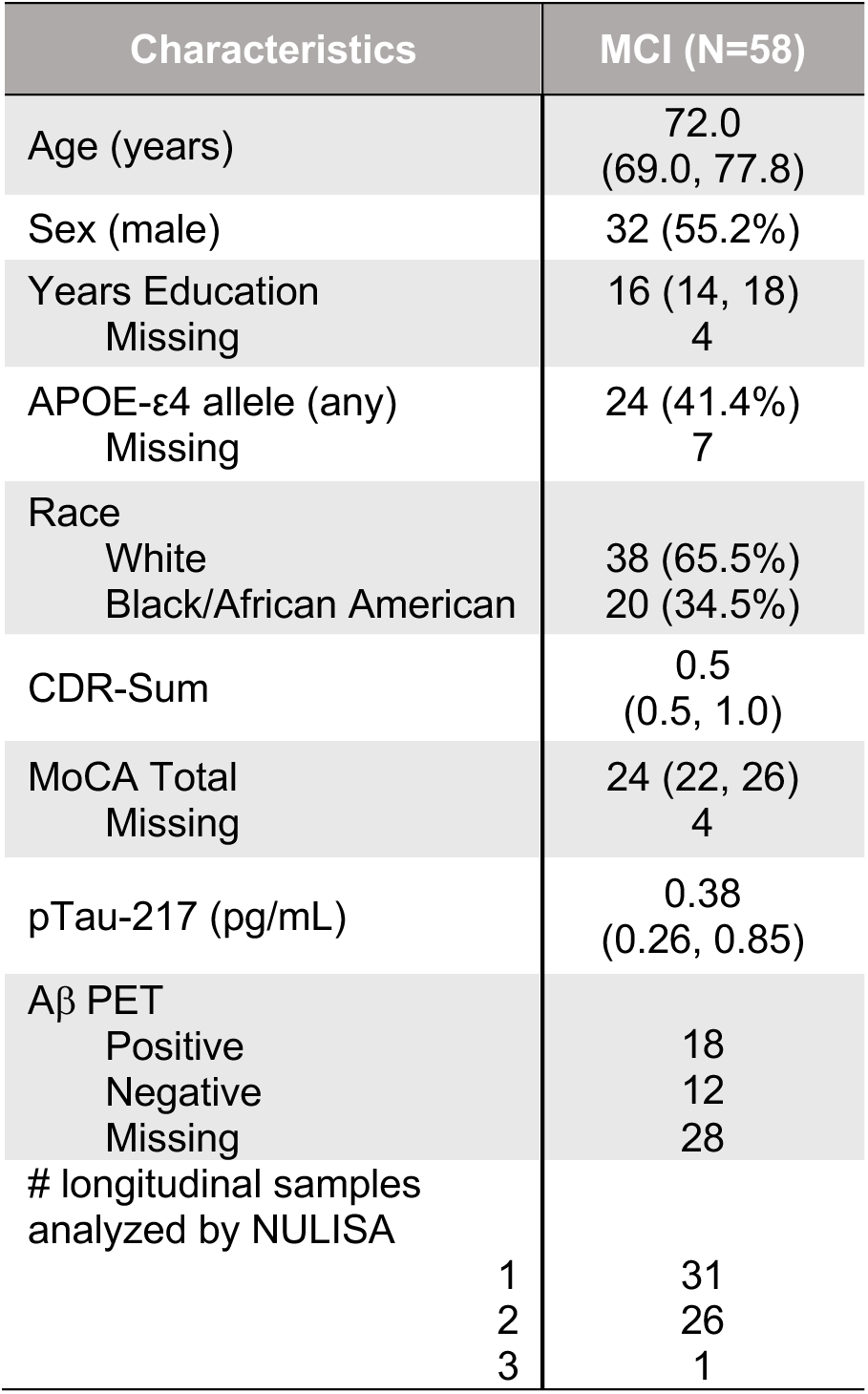
Prediction cohort of cognitively impaired individuals lacking a clear etiology. Numeric variables are summarized using median (25th percentile, 75th percentile). Categorical variables are summarized using number (percent). Abbreviations: CDR-Sum, Clinical Dementia Rating-Sum of Boxes; MoCA, Montreal Cognitive Assessment; MCI, mild cognitive impairment.

| Characteristics | MCI (N=58) |
| --- | --- |
| Age (years) | 72.0<br>(69.0, 77.8) |
| Sex (male) | 32 (55.2%) |
| Years Education<br>Missing | 16 (14, 18)<br>4 |
| APOE-ε4 allele (any)<br>Missing | 24 (41.4%)<br>7 |
| Race<br>White<br>Black/African American | 38 (65.5%)<br>20 (34.5%) |
| CDR-Sum | 0.5<br>(0.5, 1.0) |
| MoCA Total<br>Missing | 24 (22, 26)<br>4 |
| pTau-217 (pg/mL) | 0.38<br>(0.26, 0.85) |
| Aβ PET<br>Positive<br>Negative<br>Missing | 18<br>12<br>28 |
| # longitudinal samples<br>analyzed by NULISA |  |
| 1 | 31 |
| 2 | 26 |
| 3 | 1 |

Data from each individual blood sample was passed through all four calibrated models simultaneously to obtain etiology-specific probabilities (**Table S2**). Among the prediction cohort, 51.7% of the participants had a NULISA proteomic profile that was predicted to fit a dementia subtype with high probability (>0.5). This included high probability predictions of AD for 3, FTLD for 8, LBD for 10, VaD for 3 participants (**Table S2**).

### 3.6 Models including clinical and demographic variables

Adjustments for demographic and cognitive covariates can influence model-based analyses. Thus, we incorporated age, sex, years of education, CDR-Sum, and MoCA total covariates in the same models as above to enable integrated evaluation of molecular and clinical predictors. Models adjusted for these covariates achieved similar or greater discrimination between dementia subtypes (**Table 5**), and many of these covariates were strong contributors to model predictions (**Figure 6**).

**Figure 6:**
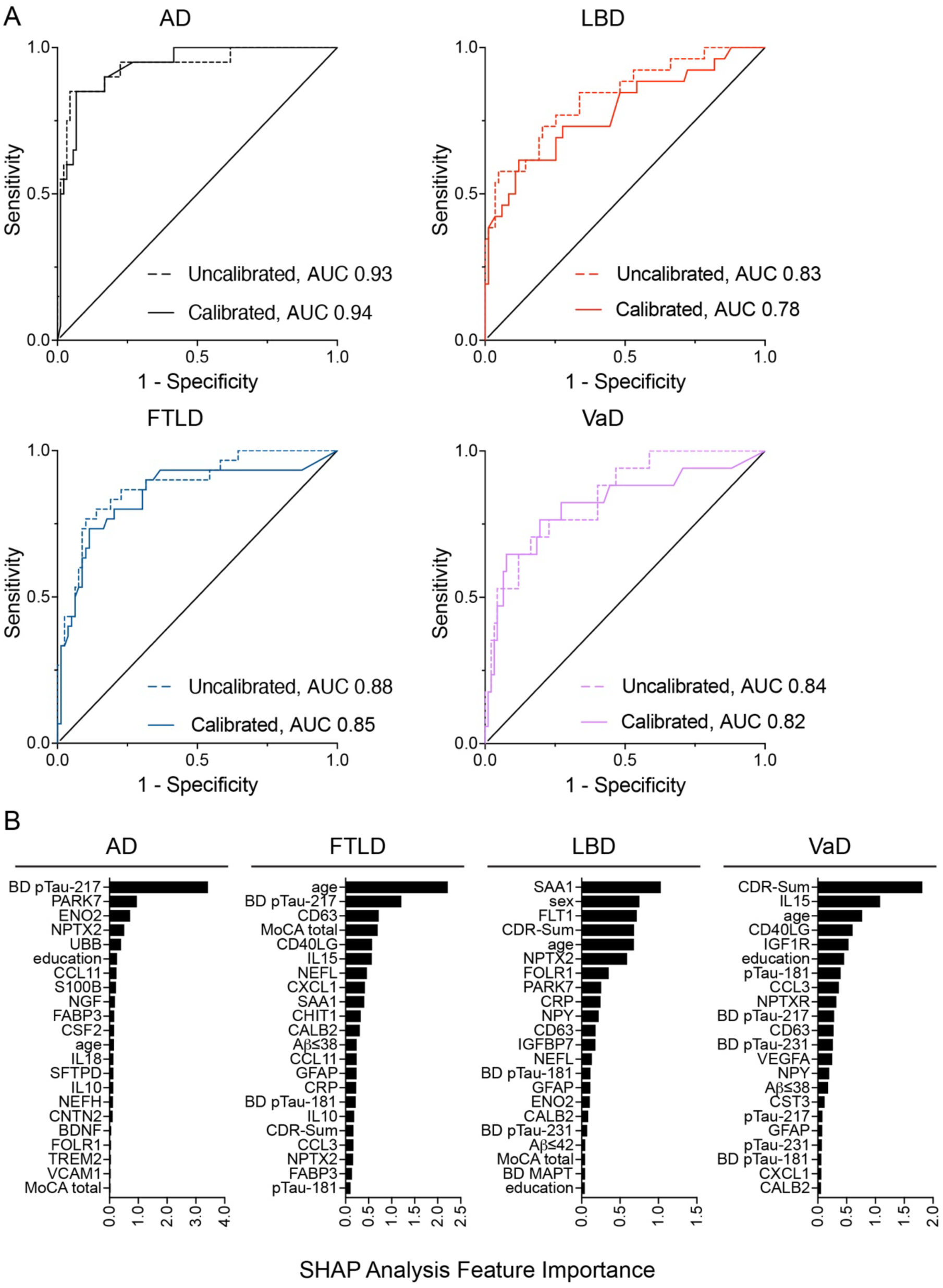
Trained XGBoost model classification discrimination and important features adjusted for age, sex, years of education, Clinical Dementia Rating-Sum of Boxes (CDR-Sum), and Montreal Cognitive Assessment total (MoCA total). (A) receiver operator characteristic curve area under the curve (AUC) for each model before (Uncalibrated) and after (Calibrated) calibration by isotonic regression comparing each participant group with the rest of the training cohort. (B) SHAP analysis of the top contributing features to each one-versus-rest model. Abbreviations: AD, Alzheimer’s disease; FTLD, frontotemporal lobar degeneration; LBD, Lewy body disease; VaD, vascular disease. Full protein names can be found in **Table S1**.

**Table 5:** Model parameters and metrics for one-versus-rest model out of fold predictions shown for each final model by participant group adjusted for age, sex, education, Clinical Dementia Rating-Sum of Boxes, and Montreal Cognitive Assessment total. XGBoost max depth of 3 was chosen for every model. Abbreviations: AD, Alzheimer’s disease; FTLD, frontotemporal lobar degeneration; LBD, Lewy body disease; VaD, vascular disease; pos, positive; neg, negative; Col sample by tree, fraction of features used for building each tree; Sens., sensitivity; Spec., specificity; ROC AUC, receiver operator characteristic curve area under the curve; Uncal., uncalibrated; Cal., calibrated; Bal., balanced.

| Group | N pos,<br>N neg | Feature p-value<br>Threshold | Col<br>Sample<br>by Tree | Learning<br>Rate | Sub-Sample | #<br>Features | N esti-<br>mators | F1<br>Score | Sens | Spec | ROC<br>AUC<br>Uncal<br>(95% CI) | ROC<br>AUC Cal<br>(95% CI) | Bal<br>Accuracy |
| --- | --- | --- | --- | --- | --- | --- | --- | --- | --- | --- | --- | --- | --- |
| AD | 20, 89 | 0.1 | 1.0 | 0.10 | 1.0 | 44 | 300 | 0.81 | 0.85 | 0.94 | 0.93<br>(0.86,<br>0.99) | 0.94<br>(0.88,<br>0.98) | 0.90 |
| FTLD | 30, 79 | 0.005 | 1.0 | 0.10 | 0.8 | 28 | 300 | 0.75 | 0.73 | 0.91 | 0.88<br>(0.79,<br>0.95) | 0.85<br>(0.74,<br>0.93) | 0.82 |
| LBD | 26, 83 | 0.01 | 1.0 | 0.03 | 0.8 | 27 | 300 | 0.67 | 0.58 | 0.95 | 0.83<br>(0.73,<br>0.93) | 0.78<br>(0.67,<br>0.88) | 0.76 |
| VaD | 17, 92 | 0.005 | 0.8 | 0.03 | 1.0 | 28 | 600 | 0.58 | 0.53 | 0.95 | 0.84<br>(0.74,<br>0.93) | 0.82<br>(0.68,<br>0.93) | 0.74 |

In the prediction cohort, 32 (55.2%) participants were predicted with high probability (>0.5) using the adjusted models compared to 30 (51.7%) using the unadjusted models (**Tables S2, S3**). The longitudinal clinical histories of twelve representative participants are shown in **Figure 7**, along with the associated model predictions. This type of modeling has the potential to detect mixed pathologies. Nine participants (10, 11, 22, 31, 40, 48, 49, 56, 58) had high probability etiology predictions for multiple dementia subtypes (**Table S3, Figure 7**). Of the 86 individual blood samples in the prediction cohort, 18 (20.9%) received a different predicted diagnosis with the covariate-adjusted models compared to the unadjusted models (**Table S4**). Only 1 of these 18 blood samples originally had a high probability diagnosis (>0.5) in the model that did not include demographic or clinical factors. That participant (participant 10, **Figure 7**) had high probability predictions of two etiologies in the unadjusted model and retained high probability predictions of the same two etiologies in the covariate-adjusted model (**Table S4**). Notably, 96.9% of the samples in the prediction cohort that had a high probability prediction using the original unadjusted models retained the same diagnostic prediction upon model adjustment for covariates, suggesting that NULISA NPQs capture the majority of sample variability introduced by these demographic and cognitive covariates (**Table S4**).

**Figure 7:**
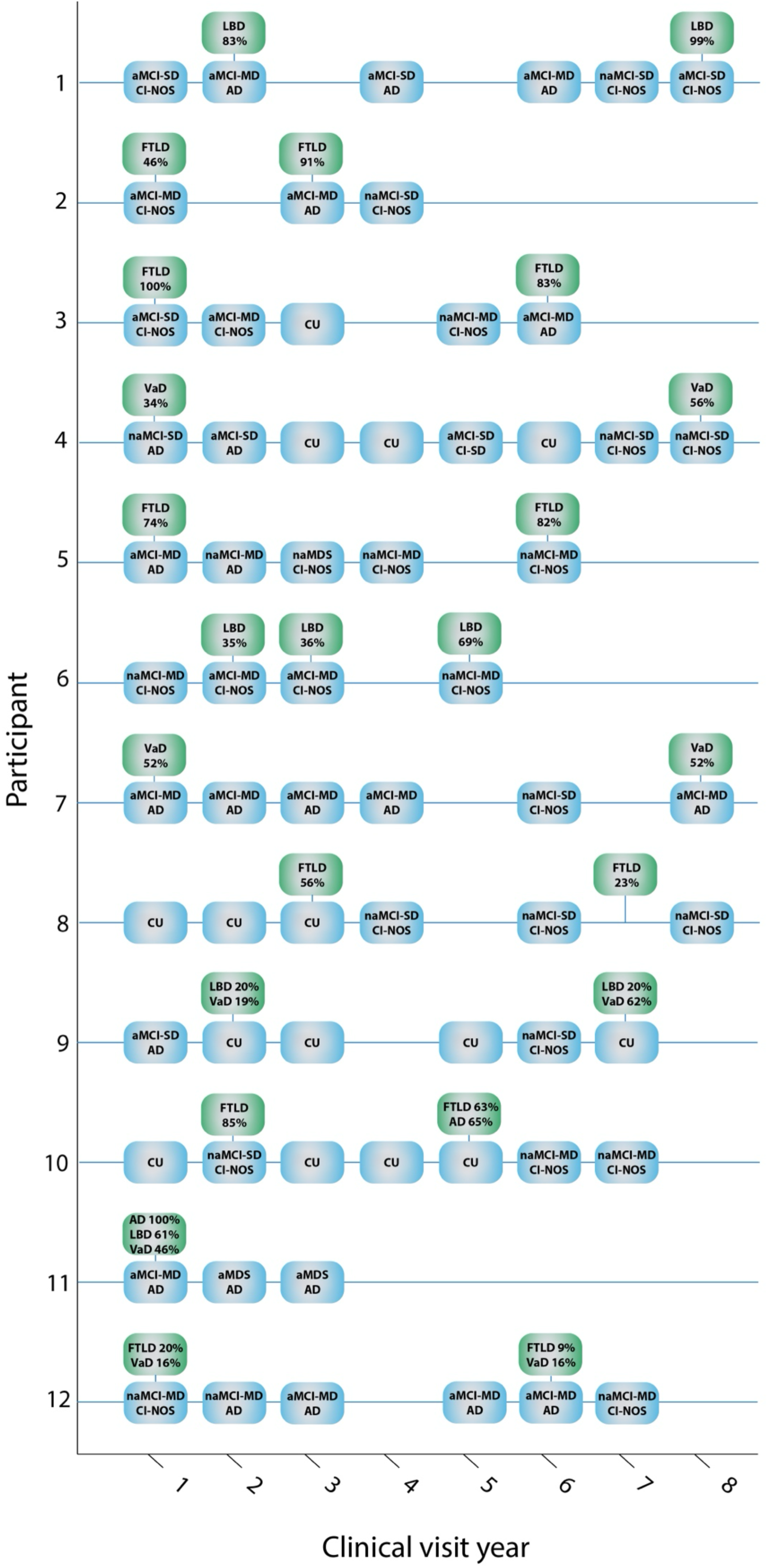
Longitudinal clinical history and model etiology predictions (from covariate-adjusted models) for participants in the prediction cohort. Longitudinal clinical diagnostic histories for twelve participants are shown across blue lines. Blue boxes represent years in which the participant was given a clinical diagnosis. The diagnosis is on the top line of each box, with the presumed etiology on the second line. Green boxes above the blue lines represent years with blood samples analyzed by NULISA. Green boxes contain the diagnostic prediction derived from the model and a % probability of that diagnosis. Abbreviations: AD, Dementia Alzheimer’s disease; FTLD, frontotemporal lobar degeneration; LBD, Lewy body disease; VaD, vascular disease; aMCI, amnestic MCI; naMCI, non-amnestic MCI; SD, single domain; MD, multiple domains; CI, cognitive impairment; NOS, not otherwise specified; aMDS, amnestic multidomain dementia syndrome; naMDS, non-amnestic multidomain dementia syndrome.

## 4. Discussion

Multiplex NULISA biomarkers have demonstrated high sensitivity for detecting changes in plasma protein levels across heterogeneous neurodegenerative disorders. Using NULISA biomarkers, we confirmed that supervised learning using XGBoost classifier models can distinguish between dementia etiologies with promising accuracy. High specificity across models indicated a tendency toward better identification of true positives rather than true negatives. Practically, this gives additional weight to high-probability predictions. In a clinical workflow, such outputs could be used to prioritize additional confirmatory testing or specialist referrals, while low-probability outputs across etiologies could flag patients for longitudinal monitoring.

Many established AD-related biomarkers, such as pTau-217, pTau-181, pTau-231, GFAP, and Aβ, were discriminatory features in these models. This finding aligns with prior evidence that NULISA biomarker measurements correlated strongly with established biomarkers of AD and related dementias using existing immunoassay platforms (such as Simoa). Our analysis of differentially abundant proteins in AD compared to CU aligns with a recent report using NULISA in a well-characterized cohort of 1596 CU and 1123 AD participants [20]. Interestingly, in our analysis, brain-derived (BD) versions of tau biomarker assays included in the NULISA CNS 120+ panel were more often important features used in classifier models than the so-called non-brain-derived versions included in the panel, demonstrating the utility of these assays to better differentiate neurodegeneration. The inclusion of inflammatory and vascular proteins (IL-6, IL-15, SAA1, VCAM1, ICAM1, CRP) underscores the contribution of neuroinflammation and vascular dysfunction to dementia. The enrichment of neuronal and stress-response proteins (PARK7, ENO2, SFTPD) in the AD-specific model highlights AD-associated neuronal stress. The importance of CHIT1 in the FTLD-specific model, along with interleukins and chemokines, points to microglial activation. The LBD-specific model relied on some features of astrocyte activation (SAA1, FLT1, GFAP). Features reflecting vascular inflammation and endothelial activation (IL-15, IGF1R, VEGFA, VCAM1) were dominant in the VaD-specific model.

Models were trained using clinically well-characterized cohorts, but the prediction cohort consisted of individuals with a clinical consensus diagnosis of MCI at least at one visit and, for some, a variable history of different diagnosed etiologies. Accordingly, we did not expect perfect concordance with profiles derived from participants with clear clinical phenotypes. The prediction cohort median CDR Sum of boxes of 0.5 underscores the relatively early disease stages represented. However, the models still yielded informative probabilities highlighting the potential for this approach to provide early clinical support for predicting etiology.

Some participants (10, 11, 22, 31, 40, 48, 49, 56, 58) were predicted to fit in multiple etiology categories with high probability. This may represent mixed or evolving pathologies. A few participants (8, 10, 25, 29, 37) with a high-probability prediction at an early blood draw had a decrease in the probability of that prediction at a later blood draw. Because of the breadth of the NULISA CNS 120+ panel, it is reasonable to suggest that these changes may be related to stage-specific biomarker fingerprints as neuroinflammation and other pathological processes changed during disease progression. For some participants (12, 14, 19, 54), consistently low probabilities across all dementia subtypes may be an indication of very early disease biology. It also may represent cognitive impairment due to etiologies not represented in our training cohort, or cognitive impairment due to non-neurodegenerative pathologies, such as sleep disturbance, medication use, or other similar factors.

Covariate-adjusted versions of the XGBoost classifiers used in this study, which incorporated key covariates known to be associated with dementia, demonstrated the power of the NULISA CNS 120+ panel in capturing sample variability related to clinical and demographic factors. Model adjustment illustrated a feasible path to further tuning phenotype prediction. Our models were not adjusted for race or Aβ PET status due to imbalanced racial representation and PET data missingness in our training cohort, but one could expect that model performance would increase if trained with a larger, more demographically balanced cohort with less missingness. While covariate-adjusted models yielded similar etiology predictions to the unadjusted models, some of the data highlight a need for additional research investigation. For example, FTLD has similar prevalence to AD under the age of 65 years [44,45] and age was the top feature affecting classification in our FTLD covariate-adjusted model, suggesting age is an important indicator of FTLD risk. In future studies, a larger, more balanced cohort will enable stronger adjustment with improved classification performance and will enable further elucidation of covariate effects.

This study represents a promising new approach for the use of NULISA plasma biomarker measurements for differential diagnosis, but there are limitations. First, the limited sample size likely affected the maximum achievable differentiation between disease etiologies. Second, postmortem pathological characterization was unavailable for the training cohort, limiting our ability to definitively confirm the etiological classification. Incorporating autopsy-confirmed cases would strengthen model training and validation in future studies. Additionally, SMOTE was used to mitigate class imbalance in the training cohort and while this is an appropriate technique for this size cohort, SMOTE may risk overfitting if not carefully cross-validated. Future work using larger, balanced cohorts will mitigate the necessity of incorporating SMOTE into the classifier models. Finally, as previously mentioned, Aβ PET status was not uniformly available for our cohort. Given the etiologic relevance of Aβ, consistent PET availability for the training cohort would likely help sharpen classification boundaries. In future studies, analysis of NULISA plasma proteomic measurements from Aβ PET positive cognitively unimpaired participants could be used to predict clinical progression or conversion to MCI over time.

Our findings demonstrated that unbiased, multiplexed plasma proteomics using NULISA offers a powerful and scalable approach to enhance diagnosis of multiple dementia etiologies. By capturing distinct protein signatures across AD, FTLD, LBD, and VaD, these biomarkers provide opportunity for distinguishing dementia subtypes, particularly in atypical or mixed presentations. Coupled with machine-learning based classification, this strategy establishes a minimally invasive framework that may enhance diagnostic confidence, improve patient stratification, and accelerate enrollment into disease-modifying therapy trials. Collectively, this work underscores the potential of NULISA plasma protein biomarkers to transform differential diagnosis in clinical and research settings, advancing the field toward precision medicine for dementia.

## Data Availability

All data produced in the present study are available upon reasonable request to the authors

## Acknowledgements

We thank the participants from the UM-MAP cohort for their time and participation. We would also like to thank the dedication and support of the MADRC staff. This work was supported by NIA P30 AG072931, R01 AG068338, R01AG058724, and the Maibach-Smiley Endowment (MSU).

## Consent Statement

All study participants provided written informed consent for these studies.

## References

[1] Ryan J, Fransquet P, Wrigglesworth J, Lacaze P. Phenotypic Heterogeneity in Dementia: A Challenge for Epidemiology and Biomarker Studies. Front Public Health 2018;6. 10.3389/fpubh.2018.00181.

[2] Armstrong RA, Lantos PL, Cairns NJ. Overlap between neurodegenerative disorders. Neuropathology 2005;25:111–24. 10.1111/j.1440-1789.2005.00605.x.

[3] 2024 Alzheimer’s disease facts and figures. Alzheimers Dement 2024;20:3708–821. 10.1002/alz.13809.

[4] Barker WW, Luis CA, Kashuba A, Luis M, Harwood DG, Loewenstein D, et al. Relative Frequencies of Alzheimer Disease, Lewy Body, Vascular and Frontotemporal Dementia, and Hippocampal Sclerosis in the State of Florida Brain Bank. Alzheimer Disease & Associated Disorders 2002;16:203.

[5] Zhang J, Zhang Y, Wang J, Xia Y, Zhang J, Chen L. Recent advances in Alzheimer’s disease: mechanisms, clinical trials and new drug development strategies. Sig Transduct Target Ther 2024;9:211. 10.1038/s41392-024-01911-3.

[6] Hering H, Bussiere T, Liu C-C, Glajch KE, Weihofen A, Grogan J, et al. A manifesto for Alzheimer’s disease drug discovery in the era of disease-modifying therapies. Molecular Neurodegeneration 2025;20:88. 10.1186/s13024-025-00872-7.

[7] Elahi FM, Miller BL. A clinicopathological approach to the diagnosis of dementia. Nat Rev Neurol 2017;13:457–76. 10.1038/nrneurol.2017.96.

[8] Jack CR, Bennett DA, Blennow K, Carrillo MC, Dunn B, Haeberlein SB, et al. NIA-AA Research Framework: Toward a biological definition of Alzheimer’s disease. Alzheimers Dement 2018;14:535–62. 10.1016/j.jalz.2018.02.018.

[9] Jack CR, Andrews JS, Beach TG, Buracchio T, Dunn B, Graf A, et al. Revised criteria for diagnosis and staging of Alzheimer’s disease: Alzheimer’s Association Workgroup. Alzheimers Dement 2024;20:5143–69. 10.1002/alz.13859.

[10] Mattsson-Carlgren N, Janelidze S, Palmqvist S, Cullen N, Svenningsson AL, Strandberg O, et al. Longitudinal plasma p-tau217 is increased in early stages of Alzheimer’s disease. Brain 2020;143:3234–41. 10.1093/brain/awaa286.

[11] Li Y, Schindler SE, Bollinger JG, Ovod V, Mawuenyega KG, Weiner MW, et al. Validation of Plasma Amyloid-β 42/40 for Detecting Alzheimer Disease Amyloid Plaques. Neurology 2022;98:e688–99. 10.1212/WNL.0000000000013211.

[12] Trelle AN, Young CB, Vossler H, Ramos Benitez J, Cody KA, Mendiola JH, et al. Plasma Aβ42/Aβ40 is sensitive to early cerebral amyloid accumulation and predicts risk of cognitive decline across the Alzheimer’s disease spectrum. Alzheimers Dement 2025;21:e14442. 10.1002/alz.14442.

[13] Pascoal TA, Aguzzoli CS, Lussier FZ, Crivelli L, Suemoto CK, Fortea J, et al. Insights into the use of biomarkers in clinical trials in Alzheimer’s disease. eBioMedicine 2024;108:105322. 10.1016/j.ebiom.2024.105322.

[14] Feng W, Beer JC, Hao Q, Ariyapala IS, Sahajan A, Komarov A, et al. NULISA: a proteomic liquid biopsy platform with attomolar sensitivity and high multiplexing. Nat Commun 2023;14:7238. 10.1038/s41467-023-42834-x.

15. NULISAseq | CNS Disease Panel. Alamar Biosciences n.d. https://alamarbio.com/nulisaseq-cns-disease-panel/ (accessed October 16, 2025).

[16] Rea Reyes RE, Wilson RE, Langhough RE, Studer RL, Jonaitis EM, Oomens JE, et al. Targeted proteomic biomarker profiling using NULISA in a cohort enriched with risk for Alzheimer’s disease and related dementias. Alzheimer’s & Dementia 2025;21:e70166. 10.1002/alz.70166.

[17] Zeng X, Lafferty TK, Sehrawat A, Chen Y, Ferreira PCL, Bellaver B, et al. Multi-analyte proteomic analysis identifies blood-based neuroinflammation, cerebrovascular and synaptic biomarkers in preclinical Alzheimer’s disease. Mol Neurodegener 2024;19:68. 10.1186/s13024-024-00753-5.

[18] Ashton NJ, Benedet AL, Molfetta GD, Pola I, Anastasi F, Fernández-Lebrero A, et al. Biomarker discovery in Alzheimer’s and neurodegenerative diseases using Nucleic Acid Linked Immuno-Sandwich Assay. Alzheimer’s & Dementia 2025;21:e14621. 10.1002/alz.14621.

[19] Wang Y-T, Ashton NJ, Therriault J, Benedet AL, Macedo AC, Pola I, et al. Identify biological Alzheimer’s disease using a novel nucleic acid–linked protein immunoassay. Brain Commun 2025;7:fcaf004. 10.1093/braincomms/fcaf004.

[20] Gong K, Timsina J, Ali M, Chen Y, Liu M, Wang C, et al. High-sensitivity plasma proteomics reveals disease-specific signatures and predictive biomarkers of Alzheimer’s disease phenotypes in a large mixed-dementia cohort. Mol Neurodegeneration 2025;20:120. 10.1186/s13024-025-00909-x.

[21] Durcan R, Heslegrave A, Swann P, Goddard J, Chouliaras L, Murley AG, et al. Novel blood-based proteomic signatures across multiple neurodegenerative diseases. Alzheimers Dement 2025;21:e70116. 10.1002/alz.70116.

[22] Gezegen H, Alaylıoğlu M, Şahin E, Swann O, Veleva E, Güven G, et al. Unravelling the plasma proteome: Pioneering biomarkers for differential dementia diagnosis. Alzheimers Dement 2025;21:e70162. 10.1002/alz.70162.

23. U-M Memory and Aging Project | University of Michigan Medical School n.d. https://medresearch.umich.edu/labs-departments/centers/madc/research/u-m-memory-and-aging-project (accessed October 16, 2025).

24. Chen T, Guestrin C. XGBoost: A Scalable Tree Boosting System. Proceedings of the 22nd ACM SIGKDD International Conference on Knowledge Discovery and Data Mining, New York, NY, USA: Association for Computing Machinery; 2016, p. 785–94. 10.1145/2939672.2939785.

25. Uniform Data Set version 3 | National Alzheimer’s Coordinating Center n.d. https://naccdata.org/data-collection/forms-documentation/uds-3 (accessed October 30, 2025).

[26] Khachaturian Z. History of Alzheimer’s Disease Research Centers: From inception in 1984 to evolution beyond 2025. Alzheimer’s & Dementia 2025;21:e70778. 10.1002/alz.70778.

[27] Morris JC. Clinical Dementia Rating: A Reliable and Valid Diagnostic and Staging Measure for Dementia of the Alzheimer Type. International Psychogeriatrics 1997;9:173–6. 10.1017/S1041610297004870.

[28] Nasreddine ZS, Phillips NA, Bédirian V, Charbonneau S, Whitehead V, Collin I, et al. The Montreal Cognitive Assessment, MoCA: A Brief Screening Tool For Mild Cognitive Impairment. Journal of the American Geriatrics Society 2005;53:695–9. 10.1111/j.1532-5415.2005.53221.x.

[29] Davis DH, Creavin ST, Yip JL, Noel-Storr AH, Brayne C, Cullum S. Montreal Cognitive Assessment for the detection of dementia. Cochrane Database Syst Rev 2021;7:CD010775. 10.1002/14651858.CD010775.pub3.

[30] Nasreddine ZS, Phillips NA, Bédirian V, Charbonneau S, Whitehead V, Collin I, et al. The Montreal Cognitive Assessment, MoCA: a brief screening tool for mild cognitive impairment. J Am Geriatr Soc 2005;53:695–9. 10.1111/j.1532-5415.2005.53221.x.

[31] Hughes CP, Berg L, Danziger WL, Coben LA, Martin RL. A new clinical scale for the staging of dementia. Br J Psychiatry 1982;140:566–72. 10.1192/bjp.140.6.566.

[32] Lynch CA, Walsh C, Blanco A, Moran M, Coen RF, Walsh JB, et al. The clinical dementia rating sum of box score in mild dementia. Dement Geriatr Cogn Disord 2006;21:40–3. 10.1159/000089218.

[33] O’Bryant SE, Lacritz LH, Hall J, Waring SC, Chan W, Khodr ZG, et al. Validation of the new interpretive guidelines for the clinical dementia rating scale sum of boxes score in the national Alzheimer’s coordinating center database. Arch Neurol 2010;67:746–9. 10.1001/archneurol.2010.115.

[34] Su Y, D’Angelo GM, Vlassenko AG, Zhou G, Snyder AZ, Marcus DS, et al. Quantitative analysis of PiB-PET with FreeSurfer ROIs. PLoS One 2013;8:e73377. 10.1371/journal.pone.0073377.

[35] Su Y, Flores S, Hornbeck RC, Speidel B, Vlassenko AG, Gordon BA, et al. Utilizing the Centiloid scale in cross-sectional and longitudinal PiB PET studies. Neuroimage Clin 2018;19:406–16. 10.1016/j.nicl.2018.04.022.

[36] Ritchie ME, Phipson B, Wu D, Hu Y, Law CW, Shi W, et al. limma powers differential expression analyses for RNA-sequencing and microarray studies. Nucleic Acids Res 2015;43:e47. 10.1093/nar/gkv007.

[37] Lemaître G, Nogueira F, Aridas CK. Imbalanced-learn: A Python Toolbox to Tackle the Curse of Imbalanced Datasets in Machine Learning. Journal of Machine Learning Research 2017;18:1–5.

[38] Chawla NV, Bowyer KW, Hall LO, Kegelmeyer WP. SMOTE: synthetic minority over-sampling technique. J Artif Int Res 2002;16:321–57.

[39] Berta E, Bach F, Jordan M. Classifier Calibration with ROC-Regularized Isotonic Regression. Proceedings of The 27th International Conference on Artificial Intelligence and Statistics, PMLR; 2024, p. 1972–80.

40. Platt J. Probabilistic Outputs for Support vector Machines and Comparisons to Regularized Likelihood Methods, 1999.

[41] Pedregosa F, Varoquaux G, Gramfort A, Michel V, Thirion B, Grisel O, et al. Scikit-learn: Machine Learning in Python. Journal of Machine Learning Research 2011;12:2825–30.

42. CalibratedClassifierCV. Scikit-Learn n.d. https://scikit-learn/stable/modules/generated/sklearn.calibration.CalibratedClassifierCV.html (accessed October 30, 2025).

[43] Lundberg SM, Erion G, Chen H, DeGrave A, Prutkin JM, Nair B, et al. From local explanations to global understanding with explainable AI for trees. Nat Mach Intell 2020;2:56–67. 10.1038/s42256-019-0138-9.

[44] Harvey RJ, Skelton-Robinson M, Rossor MN. The prevalence and causes of dementia in people under the age of 65 years. J Neurol Neurosurg Psychiatry 2003;74:1206–9. 10.1136/jnnp.74.9.1206.

[45] Ratnavalli E, Brayne C, Dawson K, Hodges JR. The prevalence of frontotemporal dementia. Neurology 2002;58:1615–21. 10.1212/wnl.58.11.1615.

