## Supplementary Materials for "Dementia etiology classification using NULISA plasma biomarkers and machine learning"

#### Table S1

| Target | Protein Name |
|---|---|
| ACHE | Acetylcholinesterase |
| AGRN | Agrin |
| ANXA5 | Annexin A5 |
| APOE | Apolipoprotein E |
| APOE4 | Apolipoprotein E |
| ARSA | Arylsulfatase A |
| Aβ38 | Amyloid-beta precursor protein |
| Aβ40 | Amyloid-beta precursor protein |
| Aβ42 | Amyloid-beta precursor protein |
| BACE1 | Beta-secretase 1 |
| BASP1 | Brain abundant membrane attached signal protein 1 |
| BD-MAPT | Brain-derived microtubule associated protein Tau |
| BD-pTau181 | Brain-derived Tau, phosphorylated at T181 |
| BD-pTau217 | Brain-derived Tau, phosphorylated at T217 |
| BD-pTau231 | Brain-derived Tau, phosphorylated at T231 |
| BDNF | Brain-derived neurotrophic factor |
| CALB2 | Calretinin |
| CCL11 | Eotaxin |
| CCL13 | C-C motif chemokine 13 |
| CCL17 | C-C motif chemokine 17 |
| CCL2 | C-C motif chemokine 2 |
| CCL22 | C-C motif chemokine 22 |
| CCL26 | C-C motif chemokine 26 |
| CCL3 | C-C motif chemokine 3 |
| CCL4 | C-C motif chemokine ligand 4 |
| CD40LG | CD40 ligand |
| CD63 | CD63 antigen |
| CHI3L1 | Chitinase-3-like protein 1; YKL-40 |

| | |
|---|---|
| CHIT1 | Chitotriosidase-1 |
| CNTN2 | Contactin-2 |
| CRH | Corticoliberin |
| CRP | C-reactive protein |
| CSF2 | Granulocyte-macrophage colony-stimulating factor |
| CST3 | Cystatin-C |
| CX3CL1 | Fractalkine |
| CXCL1 | Growth-regulated alpha protein |
| CXCL10 | C-X-C motif chemokine 10 |
| CXCL8 | Interleukin-8, IL8 |
| ENO2 | Gamma-enolase |
| FABP3 | Fatty acid-binding protein, heart |
| FCN2 | Ficolin-2 |
| FGF2 | Fibroblast growth factor 2 |
| FLT1 | Vascular endothelial growth factor receptor 1 |
| FOLR1 | Folate receptor alpha |
| GDF15 | Growth/differentiation factor 15 |
| GDI1 | Rab GDP dissociation inhibitor alpha |
| GDNF | Glial cell line-derived neurotrophic factor |
| GFAP | Glial fibrillary acidic protein |
| GOT1 | Aspartate aminotransferase, cytoplasmic |
| HBA1 | Hemoglobin subunit alpha |
| HTT | Huntingtin |
| ICAM1 | Intercellular adhesion molecule 1 |
| IFNG | Interferon gamma |
| IGF1R | Insulin-like growth factor 1 receptor |
| IGFBP7 | Insulin-like growth factor-binding protein 7 |
| IL10 | Interleukin-10 |
| IL12p70 | Interleukin-12 subunit beta_Interleukin-12 subunit alpha |
| IL13 | Interleukin-13 |
| IL15 | Interleukin-15 |
| IL16 | Pro-interleukin-16 |
| IL17A | Interleukin-17A |
| IL18 | Interleukin-18 |
| IL1B | Interleukin-1 beta |
| IL2 | Interleukin-2 |
| IL33 | Interleukin-33 |
| IL4 | Interleukin-4 |
| IL5 | Interleukin-5 |
| IL6 | Interleukin-6 |
| IL6R | Interleukin-6 receptor subunit alpha |
| IL7 | Interleukin-7 |
| IL9 | Interleukin-9 |
| KDR | Vascular endothelial growth factor receptor 2 |
| KLK6 | Kallikrein-6 |
| MAPT | Microtubule associated protein Tau |
| MDH1 | Malate dehydrogenase, cytoplasmic |
| MME | Membrane metalloendopeptidase |
| MSLN | Mesothelin |
| NEFH | Neurofilament heavy polypeptide |

| | |
|---|---|
| NEFL (or NfL) | Neurofilament light polypeptide |
| NGF | Beta-nerve growth factor |
| NPTX1 | Neuronal pentraxin-1 |
| NPTX2 | Neuronal pentraxin-2 |
| NPTXR | Neuronal pentraxin receptor |
| NPY | Neuropeptide Y |
| NRGN | Neurogranin |
| Oligo-SNCA | Alpha-synuclein |
| PARK7 | Protein/nucleic acid deglycase DJ-1 |
| PDGFRB | Platelet-derived growth factor receptor beta |
| PDLIM5 | PDZ and LIM domain 5 |
| PGF | Placenta growth factor |
| PGK1 | Phophoglycerate kinase 1 |
| POSTN | Periostin |
| PRDX6 | Peroxiredoxin-6 |
| PSEN1 | Presenilin 1 |
| pSNCA-129 | Alpha-synuclein |
| p-tau-181 | Tau, phosphorylated at T181 |
| p-tau-217 | Tau, phosphorylated at T217 |
| p-tau-231 | Tau, phosphorylated at T231 |
| pTDP43-409 | TAR DNA-binding protein 43 |
| PTN | Pleiotrophin |
| REST | RE1 silencing transcription factor |
| RUVBL2 | RuvB like AAA ATPase 2 |
| S100A12 | Protein S100-A12 |
| S100B | S100 calcium binding protein B |
| SAA1 | Serum amyloid A-1 protein |
| SFRP1 | Secreted frizzled-related protein 1 |
| SFTPD | Pulmonary surfactant-associated protein D |
| SLIT2 | Slit homolog 2 protein |
| SMOC1 | SPARC-related modular calcium-binding protein 1 |
| SNAP25 | Synaptosomal-associated protein 25 |
| SNCA | Alpha synuclein |
| SNCB | Synuclein beta |
| SOD1 | Superoxide dismutase [Cu-Zn] |
| SQSTM1 | Sequestosome-1 |
| TAFA5 | Chemokine-like protein TAFA-5 |
| TARDBP | TAR DNA-binding protein 43 |
| TEK | Angiopoietin-1 receptor |
| TIMP3 | Metalloproteinase inhibitor 3 |
| TNF | Tumor necrosis factor |
| TREM1 | Triggering receptor expressed on myeloid cells 1 |
| TREM2 | Triggering receptor expressed on myeloid cells 2 |
| UBB | Polyubiquitin |
| UCHL1 | Ubiquitin carboxyl-terminal hydrolase isozyme L1 |
| VCAM1 | Vascular cell adhesion protein 1 |
| VEGFA | Vascular endothelial growth factor A |
| VEGFD | Vascular endothelial growth factor B |
| VGF | VGF nerve growth factor inducible |
| VSNL1 | Visinin-like protein 1 |

| | |
|---|---|
| YWHAG | Tyrosine 3-monooxygenase/tryptophan 5-monoxoygenase activation protein gamma |
| YWHAZ | 14-3-3 protein zeta/delta |

**Table S1:** Protein targets included in the NULISAseq CNS Disease Panel 120.

**Figure S1**

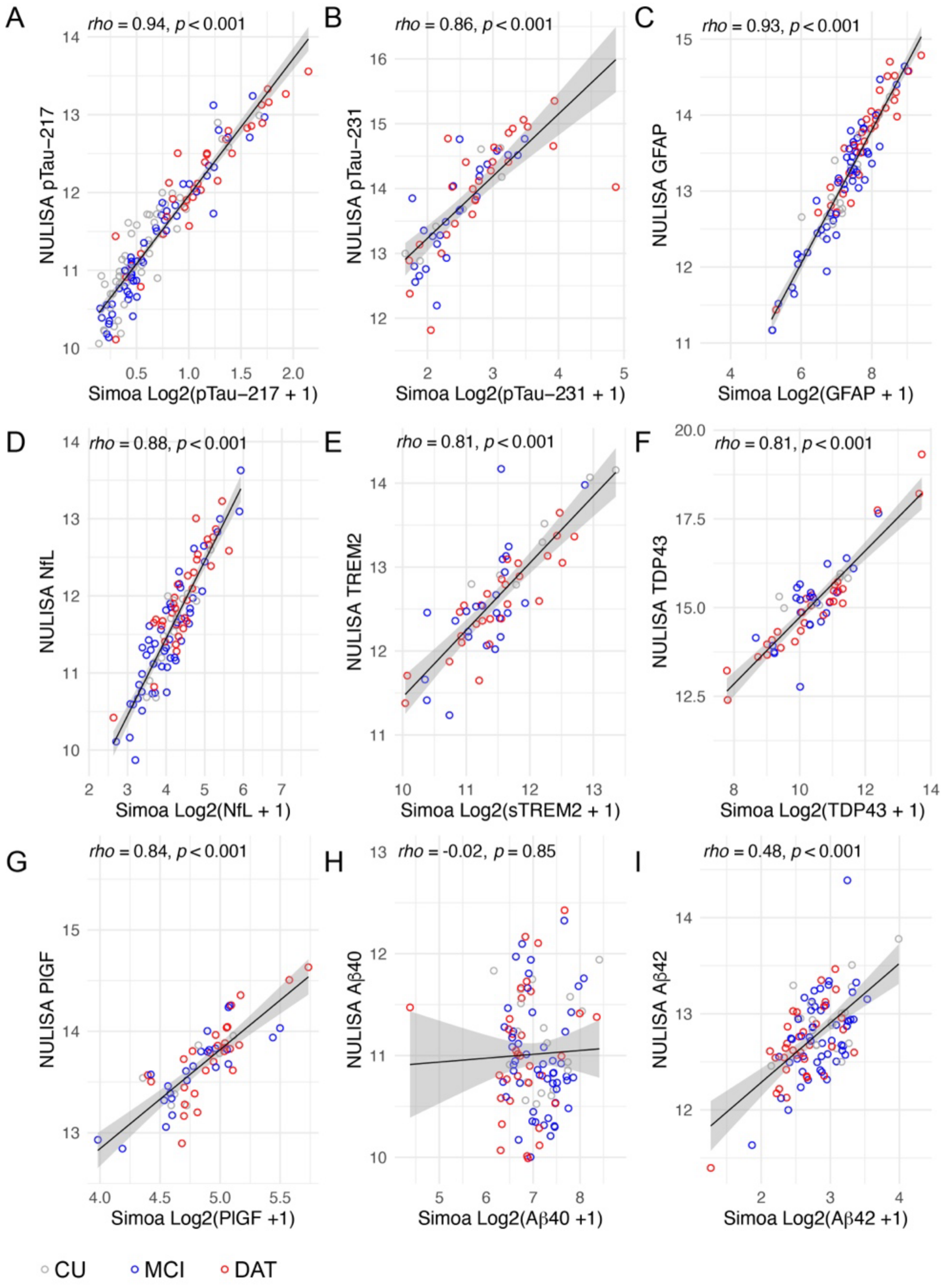


**Figure S1**: Spearman correlations between AD-related plasma biomarkers from the NULISA CNS 120+ panel and Simoa plasma biomarkers. (A) NULISA pTau-217 significantly correlates with Simoa pTau-217, rho = 0.94, $p<0.001$. (B) NULISA pTau-231 significantly correlates with Simoa pTau-231, rho = 0.86, $p<0.001$. (C) NULISA glial fibrillary acidic protein (GFAP) significantly correlates with Simoa GFAP, rho = 0.93, $p<0.001$. (D) NULISA neurofilament light chain (NfL) significantly correlates with Simoa NfL, rho = 0.88, $p<0.001$. (E) NULISA triggering receptor expressed on myeloid cells 2 (TREM2) significantly correlates with Simoa soluble TREM2, rho = 0.81, $p<0.001$. (F) NULISA Tar DNA binding protein 43 (TDP43) significantly correlates with Simoa TDP43, rho = 0.81, $p<0.001$. (G) NULISA placental growth factor (PIGF) significantly correlates with Simoa PIGF, rho = 0.84, $p<0.001$. (H) NULISA amyloid-β 40 (Aβ40) does not correlate with Simoa Aβ40, rho = -0.02, $p=0.85$. I, NULISA amyloid-β 42 (Aβ42) correlates significantly with Simoa Aβ42, rho = 0.48, $p<0.001$.

**Figure S2**

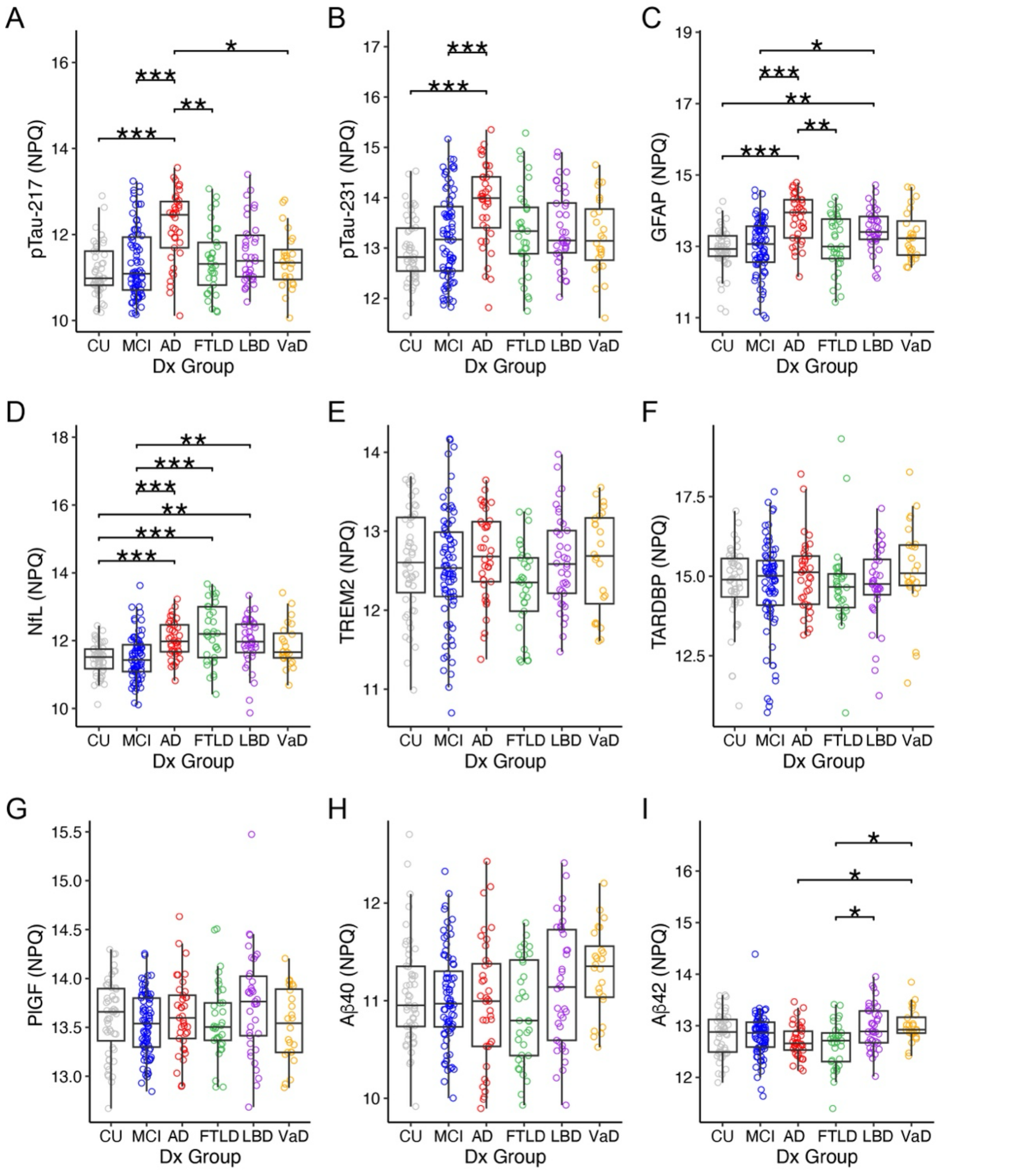


**Figure S2**: Group comparisons in the full cohort for single AD-related biomarkers from the NULISA CNS 120+ panel. (A) pTau-217 was significantly increased in AD compared to CU, MCI, FTLD, and VaD groups. (B) pTau-231 was significantly increased in AD compared to CU and MCI groups. (C) GFAP was significantly increased in AD compared to CU, MCI, and FTLD groups. It was also significantly increased in LBD compared to CU and MCI groups. (D) NfL was significantly increased in AD, FTLD, and LBD compared to CU and MCI groups. (E-H) No significant differences were observed in TREM2, TARDBP, PlGF, or Aβ40 levels among groups. (I) Aβ42 was significantly decreased in AD and FTLD compared to VaD. It was also significantly decreased in FTLD compared to LBD. Groups are compared by Kruskal-Wallis tests, p-values are from Dunn's post-hoc test. *=p<0.05, **=p<0.01, ***=p<0.001. Abbreviations: NPQ, NULISA protein quantification units; CU, cognitively unimpaired; MCI, mild cognitive impairment; AD, Alzheimer's disease; FTLD, frontotemporal lobar degeneration; LBD, Lewy body disease; VaD, vascular disease; pTau-217, phosphorylated tau at threonine 217; pTau-231, phosphorylated tau at threonine 231; GFAP, glial fibrillary acidic protein; NfL, neurofilament light chain; TREM2, triggering receptor expressed on myeloid cells 2; TARDBP, Tar DNA binding protein 43; PlGF, placental growth factor; Aβ40, amyloid-β 40; Aβ42, amyloid-β 42.

**Figure S3**

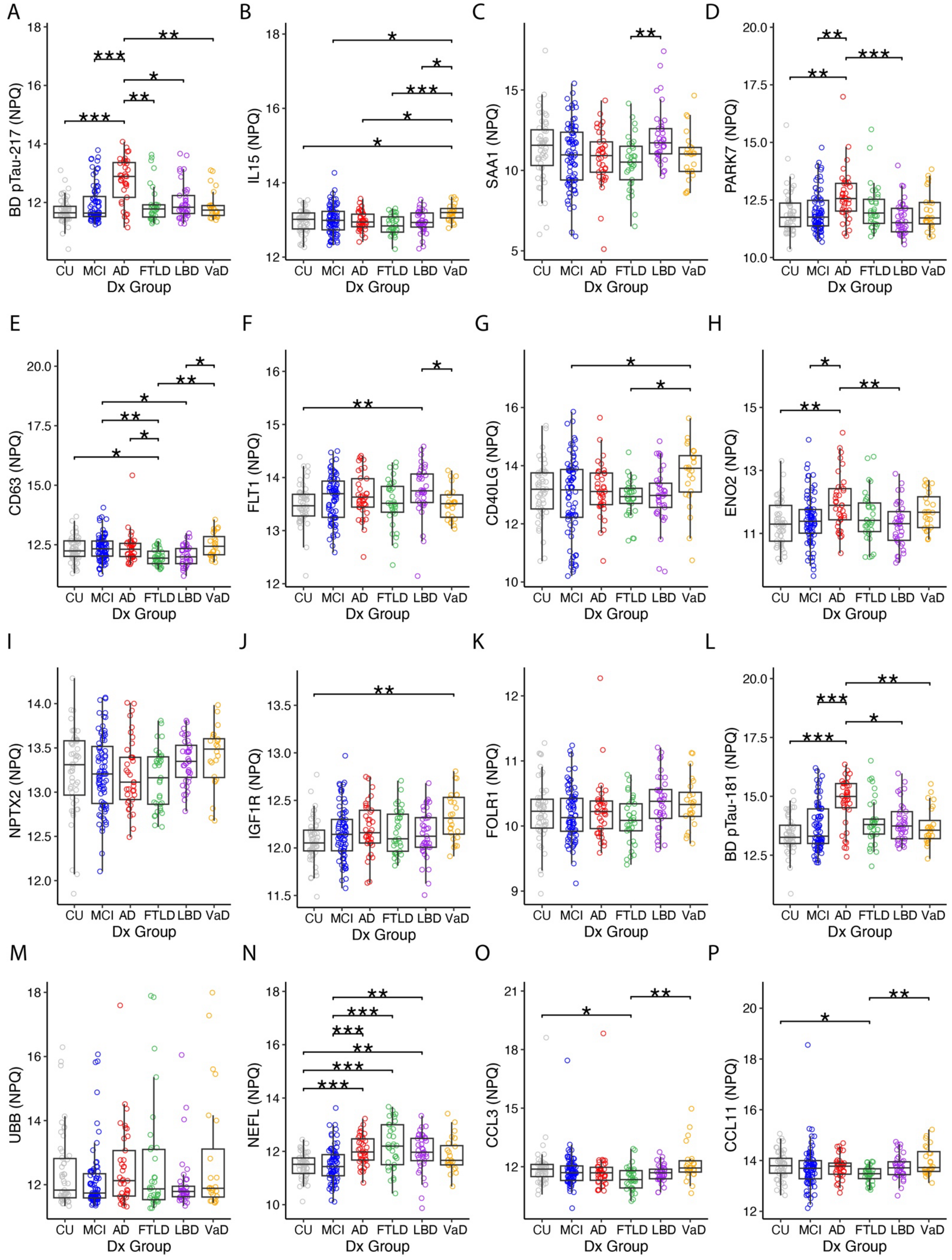
A
BD pTau-217 (NPQ)
B
IL15 (NPQ)
C
SAA1 (NPQ)
D
PARK7 (NPQ)
E
CD63 (NPQ)
F
FLT1 (NPQ)
G
CD40LG (NPQ)
H
ENO2 (NPQ)
I
NPTX2 (NPQ)
J
IGF1R (NPQ)
K
FOLR1 (NPQ)
L
BD pTau-181 (NPQ)
M
UBB (NPQ)
N
NEFL (NPQ)
O
CCL3 (NPQ)
P
CCL11 (NPQ)
CU MCI AD FTLD LBD VaD
Dx Group

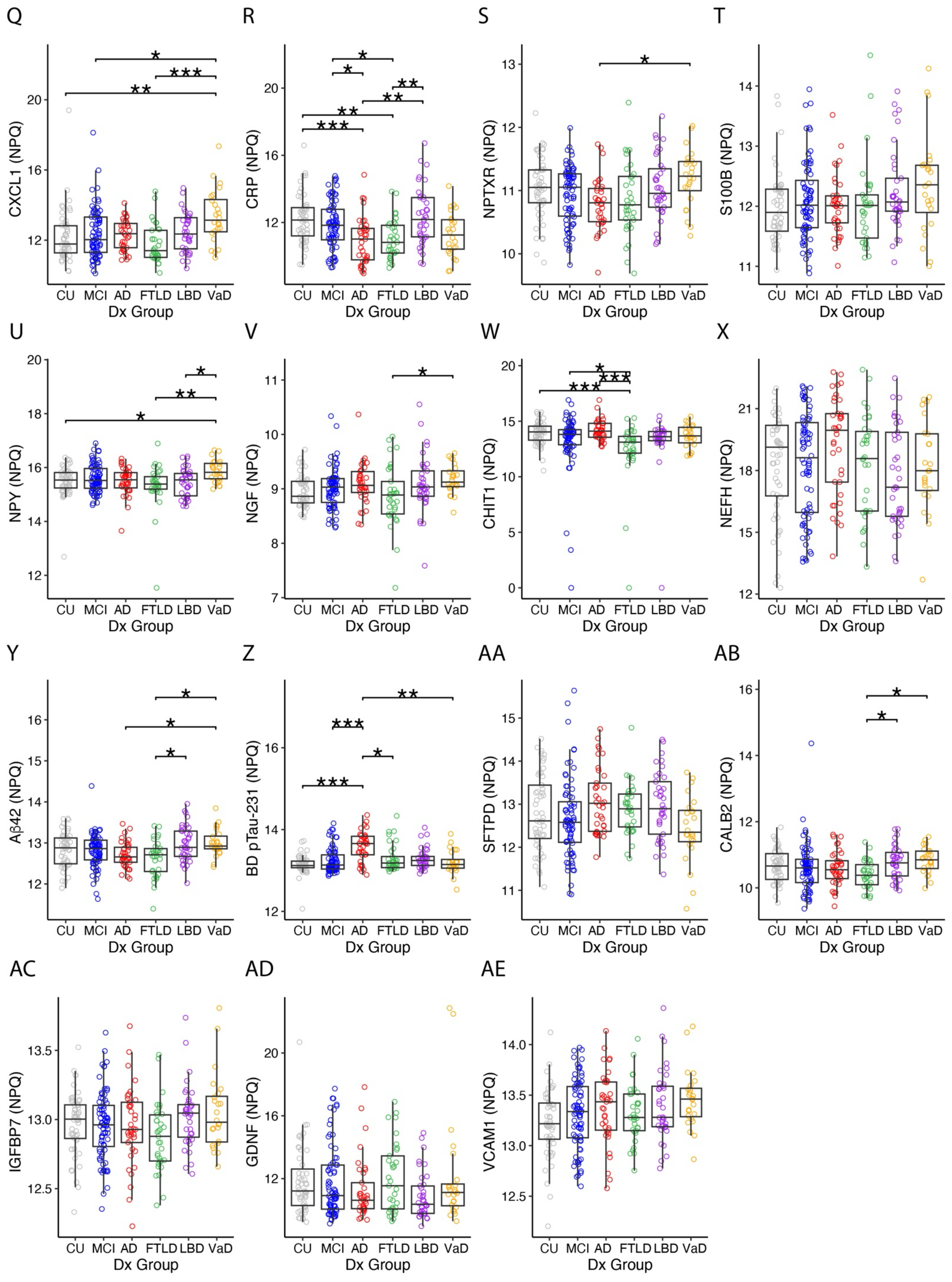
Q
CXCL1 (NPQ)
CU MCI AD FTLD LBD VaD
Dx Group
R
CRP (NPQ)
CU MCI AD FTLD LBD VaD
Dx Group
S
NPTXR (NPQ)
CU MCI AD FTLD LBD VaD
Dx Group
T
S100B (NPQ)
CU MCI AD FTLD LBD VaD
Dx Group
U
NPY (NPQ)
CU MCI AD FTLD LBD VaD
Dx Group
V
NGF (NPQ)
CU MCI AD FTLD LBD VaD
Dx Group
W
CHIT1 (NPQ)
CU MCI AD FTLD LBD VaD
Dx Group
X
NEFH (NPQ)
CU MCI AD FTLD LBD VaD
Dx Group
Y
Aβ42 (NPQ)
CU MCI AD FTLD LBD VaD
Dx Group
Z
BD pTau-231 (NPQ)
CU MCI AD FTLD LBD VaD
Dx Group
AA
SFTPD (NPQ)
CU MCI AD FTLD LBD VaD
Dx Group
AB
CALB2 (NPQ)
CU MCI AD FTLD LBD VaD
Dx Group
AC
IGFBP7 (NPQ)
CU MCI AD FTLD LBD VaD
Dx Group
AD
GDNF (NPQ)
CU MCI AD FTLD LBD VaD
Dx Group
AE
VCAM1 (NPQ)
CU MCI AD FTLD LBD VaD
Dx Group

**Figure S3**: Group comparisons from the full cohort for the top eight contributing biomarkers from the NULISA CNS 120+ panel for the unadjusted and adjusted models. The top eight biomarkers included BD pTau-217 (A), IL15 (B), SAA1 (C), PARK7 (D), CD63 (E), FLT1 (F), CD40LG (G), ENO2 (H), NPTX2 (I), IGF1R (J), FOLR1 (K), BD pTau-181 (L), UBB (M), NEFL (N), CCL3 (O), CCL11 (P), CXCL1 (Q), CRP (R), NPTXR (S), S100B (T), NPY (U), NGF (V), CHIT1 (W), NEFH (X), Aβ42 (Y), BD pTau-231 (Z), SFTPD (AA), CALB2 (AB), IGFBP7 (AC), GDNF (AD), VCAM1 (AE). Groups are compared by Kruskal-Wallis tests, *p*-values are from Dunn's post-hoc test. *=p<0.05, **=p<0.01, ***=p<0.001. See **Table S1** for full biomarker protein names.

**Table S2**

| Participant | Blood Draw Year | Aβ PET Status | Diagnostic Prediction Probability | | | |
|---|---|---|---|---|---|---|
| | | | AD | FTLD | LBD | VaD |
| 1 | 1 | unk | 0.05 | 0.05 | 0.78 | 0.23 |
| | 8 | | 0 | 0.08 | 0.92 | 0.24 |
| 2 | 1 | unk | 0.05 | 0.56 | 0.13 | 0.06 |
| | 3 | | 0 | 0.75 | 0.09 | 0.05 |
| 3 | 1 | unk | 0.05 | 0.69 | 0.04 | 0.02 |
| | 6 | | 0.07 | 0.70 | 0.15 | 0.02 |
| 4 | 1 | unk | 0 | 0.05 | 0.13 | 0.17 |
| | 8 | | 0 | 0.05 | 0.18 | 0.76 |
| 5 | 1 | unk | 0.05 | 0.61 | 0.09 | 0.02 |
| | 6 | | 0.05 | 0.61 | 0.21 | 0.02 |
| 6 | 1 | 0 | 0.06 | 0.25 | 0.27 | 0.22 |
| | 2 | | 0.46 | 0.18 | 0.78 | 0.18 |
| | 4 | | 0.11 | 0.12 | 0.78 | 0.11 |
| 7 | 1 | unk | 0.05 | 0.05 | 0.22 | 0.76 |
| | 8 | | 0.07 | 0.05 | 0.22 | 0.41 |
| 8 | 1 | unk | 0.05 | 0.65 | 0.09 | 0.02 |
| | 5 | | 0.05 | 0.30 | 0.06 | 0.11 |
| 9 | 1 | unk | 0.05 | 0.05 | 0.22 | 0.21 |
| | 6 | | 0.07 | 0 | 0.57 | 0.60 |
| 10 | 1 | unk | 0.07 | 0.86 | 0 | 0.04 |
| | 4 | | 0.84 | 0.86 | 0.11 | 0.02 |
| 11 | 1 | 1 | 0.82 | 0 | 0.58 | 0.54 |
| 12 | 1 | unk | 0.07 | 0.29 | 0.09 | 0.11 |
| | 6 | | 0.07 | 0.19 | 0.04 | 0.23 |
| 13 | 1 | unk | 0.07 | 0.33 | 0.09 | 0.11 |
| | 6 | | 0.07 | 0.05 | 0.06 | 0.22 |
| 14 | 1 | 0 | 0.07 | 0.05 | 0.04 | 0.28 |
| 15 | 1 | unk | 0.07 | 0.40 | 0.06 | 0.07 |
| | 6 | | 0 | 0.30 | 0.08 | 0.02 |
| 16 | 1 | unk | 0.07 | 0.06 | 0.04 | 0.12 |
| | 8 | | 0.07 | 0.10 | 0.04 | 0.22 |
| 17 | 1 | unk | 0.05 | 0.39 | 0.08 | 0.13 |
| | 3 | | 0.05 | 0.35 | 0.06 | 0.18 |
| 18 | 1 | 1 | 0.05 | 0.21 | 0.55 | 0.07 |
| 19 | 1 | 1 | 0.08 | 0.05 | 0.26 | 0.02 |
| 20 | 1 | unk | 0.05 | 0.18 | 0.04 | 0.11 |
| | 6 | | 0 | 0 | 0.55 | 0.11 |
| 21 | 1 | unk | 0.07 | 0.30 | 0.09 | 0.06 |

| | | | | | | |
|---|---|---|---|---|---|---|
| | 5 | | 0.07 | 0.18 | 0.10 | 0.17 |
| 22 | 1 | unk | 0.10 | 0.19 | 0.13 | 0.11 |
| | 6 | | 0.21 | 0.06 | 0.43 | 0.02 |
| 23 | 1 | unk | 0.07 | 0.24 | 0 | 0.11 |
| | 6 | | 0.07 | 0.24 | 0.04 | 0.31 |
| 24 | 1 | unk | 0 | 0.30 | 0.17 | 0.06 |
| | 4 | | 0.07 | 0.24 | 0.12 | 0.06 |
| 25 | 1 | unk | 0.05 | 0.61 | 0.10 | 0.07 |
| | 6 | | 0.05 | 0.08 | 0.01 | 0.02 |
| 26 | 1 | unk | 0.05 | 0.69 | 0.09 | 0.02 |
| 27 | 1 | 1 | 0.60 | 0.33 | 0.06 | 0.02 |
| 28 | 1 | unk | 0.07 | 0.45 | 0.06 | 0.06 |
| | 6 | | 0.07 | 0.24 | 0.08 | 0.21 |
| 29 | 1 | unk | 0.07 | 0.40 | 0.08 | 0.06 |
| | 6 | | 0.07 | 0.24 | 0 | 0.06 |
| 30 | 1 | unk | 0 | 0.19 | 0.15 | 0.15 |
| | 7 | | 0.07 | 0 | 0.56 | 0.15 |
| 31 | 1 | unk | 0.79 | 0.18 | 0.15 | 0.23 |
| 32 | 1 | 1 | 0.07 | 0.46 | 0.53 | 0.06 |
| 33 | 1 | 1 | 0.07 | 0.55 | 0.43 | 0.02 |
| 34 | 1 | 1 | 0.44 | 0.05 | 0.27 | 0.22 |
| 35 | 1 | 1 | 0.05 | 0.35 | 0.92 | 0.02 |
| 36 | 1 | 1 | 0.48 | 0.47 | 0.35 | 0.02 |
| 37 | 1 | unk | 0.05 | 0.39 | 0.29 | 0.06 |
| | 6 | | 0 | 0.12 | 0.62 | 0.22 |
| 38 | 1 | 1 | 0.84 | 0.61 | 0.15 | 0.02 |
| 39 | 1 | 1 | 0.58 | 0.05 | 0.05 | 0.21 |
| 40 | 1 | 0 | 0.07 | 0.25 | 0.68 | 0.22 |
| | 6 | | 0.07 | 0.05 | 0.81 | 0.37 |
| 41 | 1 | 1 | 0.05 | 0.30 | 0.37 | 0.13 |
| 42 | 1 | 1 | 0.05 | 0.13 | 0.22 | 0.41 |
| 43 | 1 | 0 | 0.07 | 0.38 | 0.09 | 0.18 |
| 44 | 1 | 1 | 0.40 | 0.05 | 0.78 | 0.19 |
| 45 | 1 | unk | 0.07 | 0.05 | 0.08 | 0.11 |
| 46 | 1 | 0 | 0.07 | 0.18 | 0.21 | 0.23 |
| 47 | 1 | unk | 0.07 | 0.06 | 0.08 | 0.17 |
| | 4 | | 0.07 | 0.25 | 0.27 | 0.22 |
| 48 | 1 | 0 | 0.07 | 0.08 | 0.44 | 0.68 |
| 49 | 1 | 0 | 0.07 | 0.10 | 0.48 | 0.23 |
| 50 | 1 | 0 | 0.07 | 0.08 | 0.48 | 0.22 |
| 51 | 1 | 0 | 0.07 | 0.28 | 0.06 | 0.14 |
| 52 | 1 | 0 | 0.07 | 0.03 | 0.25 | 0.18 |
| 53 | 1 | 0 | 0.07 | 0.05 | 0.42 | 0.07 |
| 54 | 1 | 0 | 0.07 | 0.35 | 0.04 | 0.15 |
| 55 | 1 | 1 | 0.07 | 0.59 | 0.30 | 0.11 |
| 56 | 1 | 1 | 0.77 | 0.05 | 0.28 | 0.75 |
| 57 | 1 | 1 | 0.49 | 0.21 | 0.09 | 0.07 |
| 58 | 1 | 1 | 0.89 | 0.05 | 0 | 0.59 |

**Table S2:** Model etiology prediction probabilities. For each longitudinal sample for each participant in the prediction cohort, a probability was generated by the model for each dementia etiology. Green, probabilities equal to or above 0.5; Yellow, probabilities between 0.25 and 0.49; Amyloid PET status,

1 = positive, 0 = negative, unk = unknown. Abbreviations: AD, Alzheimer's disease; FTLD, frontotemporal lobar degeneration; LBD, Lewy body disease; VaD, vascular disease.

**Table S3**

| Participant | Blood Draw Year | Aβ PET Status | Diagnostic Prediction Probability | | | |
|---|---|---|---|---|---|---|
| | | | AD | FTLD | LBD | VaD |
| 1 | 1 | unk | 0.03 | 0 | 0.83 | 0.17 |
| | 8 | | 0.03 | 0.09 | 0.99 | 0.56 |
| 2 | 1 | unk | 0.06 | 0.46 | 0.10 | 0.02 |
| | 3 | | 0.06 | 0.91 | 0.05 | 0.03 |
| 3 | 1 | unk | 0.03 | 1 | 0 | 0.02 |
| | 6 | | 0.06 | 0.83 | 0.18 | 0.02 |
| 4 | 1 | unk | 0.01 | 0.04 | 0.22 | 0.34 |
| | 8 | | 0 | 0.01 | 0.31 | 0.56 |
| 5 | 1 | unk | 0.04 | 0.74 | 0.06 | 0.02 |
| | 6 | | 0.03 | 0.82 | 0.20 | 0.06 |
| 6 | 1 | 0 | 0.03 | 0.35 | 0.20 | 0.40 |
| | 2 | | 0.16 | 0.35 | 0.36 | 0.29 |
| | 4 | | 0 | 0.29 | 0.69 | 0.42 |
| 7 | 1 | unk | 0.03 | 0.06 | 0.20 | 0.52 |
| | 8 | | 0.03 | 0.06 | 0.10 | 0.52 |
| 8 | 1 | unk | 0.03 | 0.56 | 0.05 | 0.08 |
| | 5 | | 0.06 | 0.23 | 0.05 | 0.06 |
| 9 | 1 | unk | 0.03 | 0.01 | 0.20 | 0.19 |
| | 6 | | 0.03 | 0 | 0.20 | 0.62 |
| 10 | 1 | unk | 0.06 | 0.85 | 0 | 0.06 |
| | 4 | | 0.65 | 0.63 | 0.06 | 0.22 |
| 11 | 1 | 1 | 1 | 0 | 0.61 | 0.46 |
| 12 | 1 | unk | 0.06 | 0.20 | 0.07 | 0.16 |
| | 6 | | 0.03 | 0.09 | 0.05 | 0.16 |
| 13 | 1 | unk | 0.06 | 0.09 | 0.20 | 0.23 |
| | 6 | | 0.03 | 0.03 | 0.47 | 0.21 |
| 14 | 1 | 0 | 0.03 | 0 | 0.05 | 0.19 |
| 15 | 1 | unk | 0.03 | 0.28 | 0.06 | 0.13 |
| | 6 | | 0.03 | 0.35 | 0.28 | 0.06 |
| 16 | 1 | unk | 0.06 | 0.35 | 0.04 | 0.16 |
| | 8 | | 0.06 | 0.17 | 0.01 | 0.41 |
| 17 | 1 | unk | 0.06 | 0.48 | 0.01 | 0.10 |
| | 3 | | 0.06 | 0.55 | 0.06 | 0.08 |
| 18 | 1 | 1 | 0.03 | 0 | 0.48 | 0.04 |
| 19 | 1 | 1 | 0.03 | 0 | 0.16 | 0.10 |
| 20 | 1 | unk | 0.03 | 0 | 0.05 | 0.18 |
| | 6 | | 0 | 0.06 | 0.76 | 0.16 |
| 21 | 1 | unk | 0.03 | 0.44 | 0.03 | 0.06 |
| | 5 | | 0.06 | 0 | 0.10 | 0.40 |
| 22 | 1 | unk | 0.03 | 0.03 | 0.10 | 0.38 |
| | 6 | | 0 | 0.11 | 0.60 | 0.71 |
| 23 | 1 | unk | 0.06 | 0.46 | 0 | 0.07 |
| | 6 | | 0.06 | 0.20 | 0.05 | 0.20 |
| 24 | 1 | unk | 0.03 | 0.35 | 0.05 | 0.24 |
| | 4 | | 0.07 | 0.11 | 0.10 | 0.26 |
| 25 | 1 | unk | 0.03 | 0.78 | 0.06 | 0.02 |
| | 6 | | 0.03 | 0.03 | 0.06 | 0.02 |

| | | | | | | |
|---|---|---|---|---|---|---|
| 26 | 1 | unk | 0.05 | 1 | 0.06 | 0.06 |
| 27 | 1 | 1 | 0.45 | 0.37 | 0 | 0.27 |
| 28 | 1 | unk | 0.03 | 0.46 | 0.05 | 0.08 |
| | 6 | | 0.06 | 0.05 | 0.05 | 0.39 |
| 29 | 1 | unk | 0.03 | 0.81 | 0.05 | 0.04 |
| | 6 | | 0.06 | 0.35 | 0 | 0.06 |
| 30 | 1 | unk | 0.03 | 0.35 | 0.18 | 0.08 |
| | 7 | | 0.03 | 0 | 0.29 | 0.08 |
| 31 | 1 | unk | 0.74 | 0 | 0.50 | 0.71 |
| 32 | 1 | 1 | 0.39 | 0.20 | 0.66 | 0.04 |
| 33 | 1 | 1 | 0.07 | 0.44 | 0.18 | 0.25 |
| 34 | 1 | 1 | 0.39 | 0.06 | 0.06 | 0.42 |
| 35 | 1 | 1 | 0.03 | 0.06 | 0.73 | 0.02 |
| 36 | 1 | 1 | 0.46 | 0.06 | 0.20 | 0.02 |
| 37 | 1 | unk | 0.06 | 0.78 | 0.10 | 0.08 |
| | 6 | | 0.03 | 0.26 | 0.70 | 0.19 |
| 38 | 1 | 1 | 0.71 | 0.35 | 0.10 | 0 |
| 39 | 1 | 1 | 0.49 | 0.06 | 0 | 0.10 |
| 40 | 1 | 0 | 0.06 | 0.06 | 0.36 | 0.36 |
| | 6 | | 0.03 | 0 | 0.83 | 0.75 |
| 41 | 1 | 1 | 0.06 | 0.20 | 0.21 | 0.38 |
| 42 | 1 | 1 | 0 | 0.20 | 0.20 | 0.59 |
| 43 | 1 | 0 | 0.06 | 0.18 | 0.31 | 0.06 |
| 44 | 1 | 1 | 0.23 | 0.06 | 0.51 | 0.46 |
| 45 | 1 | unk | 0.03 | 0.13 | 0.20 | 0.46 |
| 46 | 1 | 0 | 0.06 | 0 | 0.10 | 0.26 |
| 47 | 1 | unk | 0.03 | 0.25 | 0.05 | 0.29 |
| | 4 | | 0.06 | 0 | 0 | 0.55 |
| 48 | 1 | 0 | 0.06 | 0.06 | 0.51 | 0.81 |
| 49 | 1 | 0 | 0.07 | 0.06 | 0.61 | 0.52 |
| 50 | 1 | 0 | 0.07 | 0.06 | 0.31 | 0.29 |
| 51 | 1 | 0 | 0.06 | 0.19 | 0.28 | 0.10 |
| 52 | 1 | 0 | 0.06 | 0.11 | 0.55 | 0.33 |
| 53 | 1 | 0 | 0.06 | 0.35 | 0.28 | 0.10 |
| 54 | 1 | 0 | 0.06 | 0.19 | 0.15 | 0.11 |
| 55 | 1 | 1 | 0.06 | 0.60 | 0.10 | 0.22 |
| 56 | 1 | 1 | 0.95 | 0 | 0.28 | 0.74 |
| 57 | 1 | 1 | 0.16 | 0 | 0.10 | 0.26 |
| 58 | 1 | 1 | 0.98 | 0 | 0 | 0.75 |

**Table S3:** Model etiology prediction probabilities adjusted for age, sex, years of education, Clinical Dementia Rating-Sum of Box, and Montreal Cognitive Assessment total. Amyloid PET status is coded as 1 = positive, 0 = negative, or unk = unknown. For longitudinal samples for each participant in the prediction cohort, a probability was generated by the adjusted model for each dementia etiology. Green, probabilities equal to or above 0.5; Yellow, probabilities between 0.25 and 0.49. Abbreviations: AD, Alzheimer's disease; FTLD, frontotemporal lobar degeneration; LBD, Lewy body disease; VaD, vascular disease.

### Table S4

| Participant | Blood Draw Year | Aβ PET Status | Diagnostic Prediction: Unadjusted Model | Probability | Adjusted Model | Probability |
|---|---|---|---|---|---|---|
| 1 | 1 | unk | LBD | 0.78 | LBD | 0.83 |
| | 8 | | LBD | 0.92 | LBD | 0.99 |
| 2 | 1 | unk | FTLD | 0.56 | FTLD | 0.46 |
| | 3 | | FTLD | 0.75 | FTLD | 0.91 |
| 3 | 1 | unk | FTLD | 0.69 | FTLD | 1.00 |
| | 6 | | FTLD | 0.70 | FTLD | 0.83 |
| 4 | 1 | unk | VaD | 0.17 | VaD | 0.34 |
| | 8 | | VaD | 0.76 | VaD | 0.56 |
| 5 | 1 | unk | FTLD | 0.61 | FTLD | 0.74 |
| | 6 | | FTLD | 0.61 | FTLD | 0.82 |
| 6 | 1 | 0 | LBD | 0.27 | VaD<br>LBD | 0.40<br>0.35 |
| | 2 | | LBD | 0.78 | LBD | 0.36 |
| | 4 | | LBD | 0.68 | LBD | 0.69 |
| 7 | 1 | unk | VaD | 0.76 | VaD | 0.52 |
| | 8 | | VaD | 0.41 | VaD | 0.52 |
| 8 | 1 | unk | FTLD | 0.65 | FTLD | 0.56 |
| | 5 | | FTLD | 0.30 | FTLD | 0.23 |
| 9 | 1 | unk | LBD | 0.22 | LBD | 0.20 |
| | 6 | | VaD | 0.60 | VaD | 0.62 |
| 10 | 1 | unk | FTLD | 0.86 | FTLD | 0.85 |
| | 4 | | FTLD | 0.86 | AD<br>FTLD | 0.65<br>0.63 |
| 11 | 1 | 1 | AD | 0.82 | AD | 1.00 |
| 12 | 1 | unk | FTLD | 0.29 | FTLD | 0.20 |
| | 6 | | VaD | 0.23 | VaD | 0.16 |
| 13 | 1 | unk | FTLD | 0.33 | VaD<br>FTLD | 0.23<br>0.09 |
| | 6 | | VaD | 0.22 | LBD<br>VaD | 0.47<br>0.21 |
| 14 | 1 | 0 | VaD | 0.28 | VaD | 0.19 |
| 15 | 1 | unk | FTLD | 0.40 | FTLD | 0.28 |
| | 6 | | FTLD | 0.30 | FTLD | 0.35 |
| 16 | 1 | unk | VaD | 0.12 | FTLD<br>VaD | 0.35<br>0.16 |
| | 8 | | VaD | 0.22 | VaD | 0.41 |
| 17 | 1 | unk | FTLD | 0.39 | FTLD | 0.48 |
| | 3 | | FTLD | 0.35 | FTLD | 0.55 |
| 18 | 1 | 1 | LBD | 0.55 | LBD | 0.48 |
| 19 | 1 | 1 | LBD | 0.26 | LBD | 0.16 |
| 20 | 1 | unk | FTLD | 0.18 | VaD<br>FTLD | 0.18<br>0.03 |
| | 6 | | LBD | 0.55 | LBD | 0.76 |
| 21 | 1 | unk | FTLD | 0.30 | FTLD | 0.45 |
| | 5 | | FTLD | 0.18 | VaD<br>FTLD | 0.40<br>0 |
| 22 | 1 | unk | FTLD | 0.19 | VaD<br>FTLD | 0.38<br>0.03 |
| | 6 | | LBD | 0.43 | VaD<br>LBD | 0.71<br>0.47 |
| 23 | 1 | unk | FTLD | 0.24 | FTLD | 0.46 |

| | | | | | | |
|---|---|---|---|---|---|---|
| | 6 | | VaD | 0.31 | VaD | 0.20 |
| 24 | 1 | unk | FTLD | 0.30 | FTLD | 0.33 |
| | 4 | | FTLD | 0.24 | VaD<br>FTLD | 0.26<br>0.35 |
| 25 | 1 | unk | FTLD | 0.61 | FTLD | 0.78 |
| | 6 | | FTLD | 0.08 | LBD<br>FTLD | 0.06<br>0.03 |
| 26 | 1 | unk | FTLD | 0.69 | FTLD | 1 |
| 27 | 1 | 1 | AD | 0.60 | AD | 0.45 |
| 28 | 1 | unk | FTLD | 0.45 | FTLD | 0.46 |
| | 6 | | FTLD | 0.24 | VaD<br>FTLD | 0.39<br>0.05 |
| 29 | 1 | unk | FTLD | 0.40 | FTLD | 0.81 |
| | 6 | | FTLD | 0.24 | FTLD | 0.35 |
| 30 | 1 | unk | FTLD | 0.19 | FTLD | 0.35 |
| | 7 | | LBD | 0.56 | LBD | 0.29 |
| 31 | 1 | unk | AD | 0.79 | AD | 0.74 |
| 32 | 1 | 1 | LBD | 0.53 | LBD | 0.66 |
| 33 | 1 | 1 | FTLD | 0.56 | FTLD | 0.44 |
| 34 | 1 | 1 | AD | 0.44 | VaD<br>AD | 0.42<br>0.39 |
| 35 | 1 | 1 | LBD | 0.92 | LBD | 0.73 |
| 36 | 1 | 1 | AD | 0.48 | AD | 0.46 |
| 37 | 1 | unk | FTLD | 0.39 | FTLD | 0.78 |
| | 6 | | LBD | 0.62 | LBD | 0.70 |
| 38 | 1 | 1 | AD | 0.84 | AD | 0.71 |
| 39 | 1 | 1 | AD | 0.58 | AD | 0.49 |
| 40 | 1 | 0 | LBD | 0.68 | LBD | 0.36 |
| | 6 | | LBD | 0.81 | LBD | 0.83 |
| 41 | 1 | 1 | LBD | 0.37 | VaD<br>LBD | 0.38<br>0.21 |
| 42 | 1 | 1 | VaD | 0.41 | VaD | 0.59 |
| 43 | 1 | 0 | FTLD | 0.38 | LBD<br>FTLD | 0.31<br>0.18 |
| 44 | 1 | 1 | LBD | 0.78 | LBD | 0.51 |
| 45 | 1 | unk | VaD | 0.11 | VaD | 0.46 |
| 46 | 1 | 0 | VaD | 0.23 | VaD | 0.26 |
| 47 | 1 | unk | VaD | 0.17 | VaD | 0.29 |
| | 4 | | LBD | 0.27 | VaD<br>LBD | 0.55<br>0 |
| 48 | 1 | 0 | VaD | 0.68 | VaD | 0.81 |
| 49 | 1 | 0 | LBD | 0.48 | LBD | 0.61 |
| 50 | 1 | 0 | LBD | 0.48 | LBD | 0.31 |
| 51 | 1 | 0 | FTLD | 0.28 | LBD<br>FTLD | 0.28<br>0.19 |
| 52 | 1 | 0 | LBD | 0.25 | LBD | 0.55 |
| 53 | 1 | 0 | LBD | 0.42 | FTLD<br>LBD | 0.35<br>0.28 |
| 54 | 1 | 0 | FTLD | 0.35 | FTLD | 0.19 |
| 55 | 1 | 1 | FTLD | 0.59 | FTLD | 0.60 |
| 56 | 1 | 1 | AD | 0.77 | AD | 0.95 |
| 57 | 1 | 1 | AD | 0.49 | VaD<br>AD | 0.26<br>0.16 |
| 58 | 1 | 1 | AD | 0.89 | AD | 0.98 |

**Table S4:** Dementia etiology predicted probabilities for unadjusted models and models adjusted for age, sex, years of education, Clinical Dementia Rating-Sum of Boxes, and Montreal Cognitive Assessment total. Amyloid PET status is coded as 1 = positive, 0 = negative, or unk = unknown. For each longitudinal sample for each participant in the prediction cohort, etiology prediction and probability is shown for the unadjusted models and the adjusted models. Yellow, etiology predictions that were different in the adjusted models compared to the unadjusted models. For these samples, the probability of the original prediction is included below the adjusted prediction. Abbreviations: AD, Alzheimer's disease; FTLD, frontotemporal lobar degeneration; LBD, Lewy body disease; VaD, vascular disease; CDR-Sum, Clinical Dementia Rating-Sum of Boxes; MoCA, Montreal Cognitive Assessment.
